# Genomic Architecture of Migraine: A Multi-ancestry GWAS Meta-analysis of 2.5 Million Participants

**DOI:** 10.64898/2026.08.28.26361638

**Authors:** Cassie Overstreet, Marco Galimberti, Kazi Tanvir Hasan, Sarah Beck, Jack Hirsch, Sanjeev Sariya, Brian Ferolito, Yu Zhou, Yingzhe Zhang, Ellen I. Weinheimer, AnnMarie Lacobelle, Yaira Nunez, The VA Million Veteran Program, Henry R. Kranzler, J. Michael Gaziano, Murray B. Stein, Christopher Gottschalk, Karmel Choi, Alexandre C. Pereira, Joseph D. Deak, Gita A Pathak, Daniel F. Levey, Joel Gelernter

## Abstract

Migraine is a leading cause of disability^1^, yet preventive treatment remains largely empirical despite the availability of several mechanistically distinct therapies^2^. Genetic data can clarify mechanisms and therapeutic hypotheses when association signals are integrated with molecular and clinical data^3^. We meta-analyzed migraine GWAS data from 12 European ancestry cohorts (206,893 cases and 2,093,175 controls) and four African ancestry cohorts (22,115 cases and 178,626 controls). We identified 311 lead variants in European-ancestry analyses and 316 lead variants in trans-ancestry analysis. Fine-mapping and transcriptome-wide analyses prioritized variants and genes implicated in sensory neuronal signaling, vascular tone, and immune regulation, with convergent evidence at several established loci including *TRPM8* and *PHACTR1*^4^. Drug-repurposing analyses identified therapeutic targets and compounds, including established migraine treatments and candidates requiring experimental validation. Genetic correlations, Mendelian randomization, and a phenome-wide scan linked migraine liability to psychiatric, pain, and gastrointestinal phenotypes. Together, these findings expand the known genetic architecture of migraine across ancestries and provide a genetics-led map connecting association signals with biological pathways, multimorbidity and candidate therapeutic mechanisms, providing a foundation for future functional and translational studies.

## Introduction

Migraine is a highly prevalent neurovascular disorder, with an estimated lifetime prevalence of 14–15% globally^5^. It is characterized by recurrent episodes of moderate-to-severe pain often accompanied by sensitivity to light and sound and odors, visual disturbances, nausea or vomiting, affective disturbance (e.g. anxiety, euphoria or depression) and cognitive impairment (impaired attention and recall, often subsumed under “brain fog”)^6^. Many are severely affected, with migraine representing one of the leading causes of disability worldwide (second overall across all ages and both sexes, and first among young women^1^). Migraine is frequently comorbid with a range of physical^7^ and psychiatric disorders^8^. Despite the availability of acute pharmacotherapies such as triptans and preventive agents ranging from anti-seizure medications to calcitonin gene-related peptide (CGRP) antagonists^9^, migraine treatment frequently involves substantial trial and error to identify effective treatment strategies and avoid common medication concerns such as acute medication overuse and potential adverse effects (e.g., drug interactions between migraine medications and treatments for comorbid conditions)^10^.

Genetic factors play a significant role in migraine etiology, with heritability estimates from twin studies ranging between 35-60%^11^. The most recent large-scale genome-wide association study (GWAS) of migraine in European (EUR) ancestry populations^4^ included 102,084 migraine cases (defined via clinical interview and self-report) and identified 123 genome-wide significant (GWS) loci. The strongest associations, mapping to genes including *LRP1*, *PHACTR1*, and *TRPM8,* have been linked to biological pathways associated with migraine pathophysiology, including glutamatergic neurotransmission, vascular tone regulation, ion channel function, and sensory neuronal signaling^12^. A subsequent meta-analysis^13^, which incorporated some of the same biobank’s datasets, demonstrated that rare variants contribute substantially in addition to the common variants highlighted previously^4^. Additional research is warranted to refine our understanding of migraine-relevant biological pathways and the underlying genetic architecture, particularly regarding treatment implications. Genetically informed drug-repurposing may offer a useful strategy of overcoming novel drug trial limitations, reducing the time and expense associated with new drug approvals.

To further refine our understanding of underlying migraine biology, larger sample sizes are needed for GWAS and post-GWAS analyses (e.g., Mendelian randomization and drug-repurposing analyses). Therefore, we performed GWAS analyses and meta-analyses of migraine in European (EUR; 206,893 migraine cases and 2,093,175 controls) and African (AFR; 22,115 cases and 178,626 controls) ancestry participants, conducting: (1) meta-analyses of GWAS results across six GWAS datasets derived from 12 underlying cohorts including novel GWAS performed herein (All of Us [AoU^14^], Mount Sinai Million Health Discoveries Program [MSMHD^15^], Yale-Penn [YP^16^]), (2) post-GWAS in-silico analyses, including genetic correlations, transcriptome-wide association analyses, Mendelian randomization, fine mapping, phenome-wide association study, and drug-repurposing analyses, and (3) residual-based DNA methylation aging as it relates to migraine case status in the MVP cohort using the Horvath epigenetic clock method^17^. Building upon prior large-scale migraine genetics studies, this work increases sample size and applies multiple complementary analytic approaches to further characterize the genetic architecture of migraine and its treatment.

## Results

### Phenotypic Harmonization, SNP-based Heritability Estimates, and Genetic Correlations between Cohorts

PheMED^30^ was performed to assess potential bias associated with phenotypic dilution (e.g., decreased power due to possible case/control misclassification; Supplementary Table 1). PheMED estimates (φ*MED*) for EUR cohorts ranged from 1.047 (CI=0.993-1.101, *p*=.085; AoU) to 1.259 (CI=1.196-1.331, *p*≈1×10⁻¹⁷; MVP) indicating phenotypic dilution ranging from non-significant minimal dilution to significant minimal dilution, respectively. For AFR cohorts, φ*MED* of 1.499 (CI=1.147-2.067; *p*=5.7×10^−11^) was identified between MVP and AoU, indicating statistically significant moderate phenotypic dilution. YP and MSMHD were not included in the PheMED analyses due to lack of power reflected in non-significant *h²_SNP_* estimates.

Liability-scale *h²_SNP_* estimates ranged from 0.085 (SE=0.004; 2023 meta-analysis) to 0.118 (SE=0.006; 23andMe). Genetic correlations across EUR cohorts ranged from 0.867 (SE=0.039; MVP-AoU) to 0.997 (SE=0.025; 23andMe-2023 meta-analysis). Additional details are presented in Supplementary Table 2. LDSC produced non-significant, near-zero observed *h²_SNP_* estimates for the MSMHD and YP EUR cohorts; therefore, genetic correlations were not calculated.

For AFR cohorts, the liability-scale *h²_SNP_* estimate for MVP was 0.083 (SE=0.024) and 0.039 (SE=0.027) for AoU. The genetic correlation between MVP-AoU cohorts was *r_g_*=0.790, SE=0.397 (*p*=0.047). As in EUR, MSMHD and YP AFR cohorts had non-significant *h*²*_SNP_* estimates and were therefore not included in genetic correlation analyses.

The high genetic correlations across EUR cohorts, in addition to the PheMED results, indicated that meta-analysis would be appropriate for EUR (i.e., the cohorts did not differ substantially genetically, and only minimal phenotypic dilution was present).

Using Popcorn to derive trans-ancestry genetic correlations between EUR and AFR meta-analysis results, we identified significant liability-scale SNP-based heritability estimates for both EUR (*h²_SNP_*=0.074, *p*=4.53×10^−80^) and AFR (*h²_SNP_*=0.051, *p*=1.02×10^−4^) ancestries. Differences in *h²_SNP_* between LDSC and Popcorn likely reflect differences in LD reference panels, SNP sets, and models utilized (i.e., univariate for LDSC and bivariate for Popcorn). The Popcorn results demonstrated that the correlation of effect sizes weighted by allele frequency across ancestries was significantly different from 1 (PGI=0.549, SE=0.082, *p*=4.03×10^−8^) indicating moderately shared genetic architecture between EUR and AFR. We additionally observed that the correlation of variant effects across ancestries when allele frequency was not considered was significantly different from 1 (PGE=0.648, SE=0.102, *p*=5.79×10^−4^), further supporting partially shared but distinct genetic architectures across ancestries.

### European Ancestry Meta-analysis

Meta-analysis of EUR ancestry cohorts identified 233 genomic risk loci, 311 lead variants (Supplementary Table 3), and 834 independent significant SNPs (Figure 1). The LDSC intercept was 1.048 (SE=0.012) and the liability-scale *h²_SNP_* estimate was 0.087 (SE=0.003).

**Figure 1.**
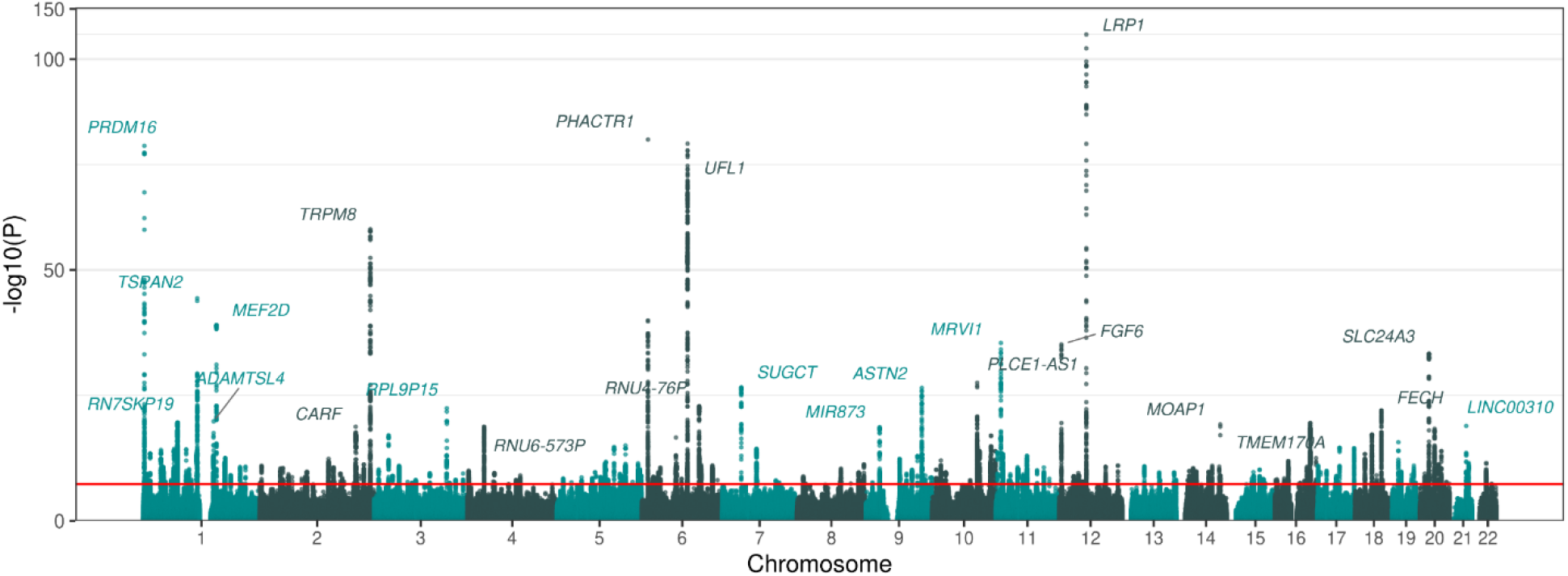
Manhattan plot of the European ancestry migraine genome-wide association meta-analysis. The red dashed line indicates the genome-wide significance threshold (*p* = 5×10^-8^).

The *r_g_* between the EUR meta-analysis *excluding* 23andMe and the 23andMe dataset alone was 0.977 (SE=0.020). Meta-analysis *excluding* 23andMe revealed 121 genomic risk loci, 159 lead variants, and 409 independent significant SNPs (Supplementary Table 4; Extended Figure 1). The LDSC intercept was 1.043 (SE=0.010) and the liability-scale *h²_SNP_* estimate was 0.083 (SE=0.003).

### GWAS Results in Novel Samples

Three novel cohorts were included in the meta-analysis described above, here we summarize individual cohort results. In the AoU EUR GWAS (*N*_eff_=143,383.4), 9 genomic risk loci, 19 independent GWS variants, and 9 lead variants were identified (Supplementary Table 5; Extended Figure 2). The variant with the smallest *p*-value was *LRP1\**rs11172113 (β=-0.072, SE=0.008, *p*=2.744×10^−19^). One lead variant, *LINC00698\**rs72887526, was identified in the AoU AFR GWAS (*N*_eff_=28,825.9, β=-0.111, SE=0.020, *p*=1.97×10^−8^; Supplementary Table 5; Extended Figure 3). No GWS variants were identified in the GWAS performed in MSMHD in either EUR (*N*_eff_=4,897.2) or AFR (*N*_eff_=4,984.5). Finally, GWAS were performed for each of the three YP cohorts and then meta-analyzed. Two lead variants were identified in the YP EUR meta-analysis (*N*_eff_=2,786.7, *AMIGO1\**rs2781555, β=-0.055, SE=0.010, *p*=2.31×10^−8^ and *BEND7\**rs7096196, β=-0.052, SE=0.009, *p*=3.83×10^−8^; Supplementary Table 6; Extended Figure 4). No GWS variants were identified in the YP AFR meta-analysis (*N*_eff_=2,862.7; Extended Figure 5).

### African Ancestry Meta-analysis

There were two GWS variants in the meta-analysis across AFR ancestry cohorts (Supplementary Table 7; Extended Figure 6), *AC019055.1\**rs10188451 (β=0.071, SE=0.015, *p*=2.57×10^−8^) and *AP1S3\**rs146372138 (β=0.132, SE=0.026, *p*=2.70×10^−9^). The LDSC intercept was 1.038 (SE=0.009). The AFR ancestry migraine meta-analysis liability-scale *h²_SNP_* estimate was 0.059 (SE=0.014, *p*=1.37×10^−5^). Neither variant, nor any LD proxy identified in the AFR migraine meta-analysis was present in the EUR meta-analysis results. Both rs10188451 and rs146372138 were polymorphic in AFR reference panels (MAF=0.037 and 0.013, respectively) but showed no observed variation in EUR.

### Trans-ancestry Meta-analysis

The trans-ancestry meta-analysis of EUR and AFR migraine meta-analyses revealed 200 genomic risk loci, 316 lead variants (Supplementary Table 8), and 981 independent significant SNPs. The corresponding Manhattan plot (EUR and AFR) is presented in Extended Figure 7.

### Gene-based Analyses

MAGMA gene-based analyses indicated tissue-specific enrichment in cervix, uterus, heart, brain, and pancreas (Extended Figure 8). MAGMA gene-set analyses also showed substantial overlap with previously reported GWAS Catalog^36^ associations across multiple domains, including psychiatric disorders (schizophrenia [*p*_adj_=5.92×10^−3^^9^], bipolar disorder [*p*_adj_=1.59×10^−2^^9^], anorexia nervosa [*p*_adj_=4.25×10^−8^]), broadly-defined headache (*p*_adj_ =1.81×10^−1^^5^), chronic obstructive pulmonary disease or coronary artery disease (*p*_adj_=6.52×10^−1^^5^) and other traits including inflammatory diseases, dietary factors, and body weight distribution traits presented in Supplementary Table 8.

### Comparisons with Prior Analyses

To quantify the degree of shared genetic signal between our present EUR migraine meta-analysis and the most recent prior common variant meta-analysis (2022; ∼63,990 overlapping participants)^4^, we compared LD-independent lead SNPs from each study and then assessed whether nonidentical SNPs were in high LD, to distinguish truly independent signals from those tagging the same association. The most recent prior meta-analysis^4^ identified 123 lead SNPs compared to 311 in the present meta-analysis. Loci identified in our study were defined using FUMA default lead-SNP and ±250 kb window approaches, which differs from the custom LD-based pruning strategy used in the prior study^4^ (iterative *p*-value ranked pruning at *r²*>0.10 followed by broadening high-LD blocks to *r²*>0.60 and merging within ±250 kb). Twenty identical variants were shared across lists and required no LD comparisons. Next, we computed pairwise LD between all remaining SNPs (103 in the prior, 291 in the present) in the two lists using a shared reference panel (1KG EUR^18^) and identified all SNP pairs with high LD (r²≥0.40) across meta-analysis results lists. In total, 62 lead variants from the present meta-analysis were in high LD with lead SNPs identified in the previous meta-analysis^4^, leaving 229 putatively novel lead variants from our present study (Supplementary Table 10) and evidence supporting 82 of the 123 previously reported variants.

### Fine Mapping

Using echolocatoR in EUR, we identified 24 high-confidence consensus SNPs that were included in the 95% credible sets of both SuSiE and FINEMAP and had a mean PP≥0.95 (Supplementary Table 11). Multiple variants mapped to genes that have been previously linked with migraine (e.g., *TRPM8, PHACTR1, STAT6, SLC24A3, RNF213*); however, less frequently discussed genes identified through fine mapping included *MRVI1* [*IRAG1*], *CARF*, and *ENDOV*.

### Genetic Correlations

A total of 900 significant associations were identified across multiple phenotypic domains (Figure 2, Supplementary Table 12) using FDR-adjusted *p*<0.05. Traits reflecting common migraine symptoms that were identified included: visual disturbances (*r_g_*=0.479, SE=0.040, FDR-adjusted *p*=5.22×10^-31^, consistent with migraine with aura) and nausea and vomiting (*r_g_*=0.454, SE=0.029, FDR-adjusted *p*=1.99×10^-54^, also consistent with common migraine symptoms). Other notable associations included other headache syndromes (*r_g_*=0.737, SE=0.024, FDR-adjusted *p*=1.37×10^−201^), gastroesophageal reflux disease (GERD; *r_g_*=0.414, SE=0.020, FDR-adjusted *p*=1.00×10^−9^^1^), chronic pain (*r_g_*=0.472, SE=0.027, FDR-adjusted *p*=8.98×10^−6^^8^), opioid use disorder (*r_g_*=0.121, SE=0.046, FDR-adjusted *p*=1.81×10^−2^), and serum potassium levels (minimum per labs; *r_g_*=-0.241, SE=0.019, FDR-adjusted *p*=1.00×10^−3^^4^).

**Figure 2.**
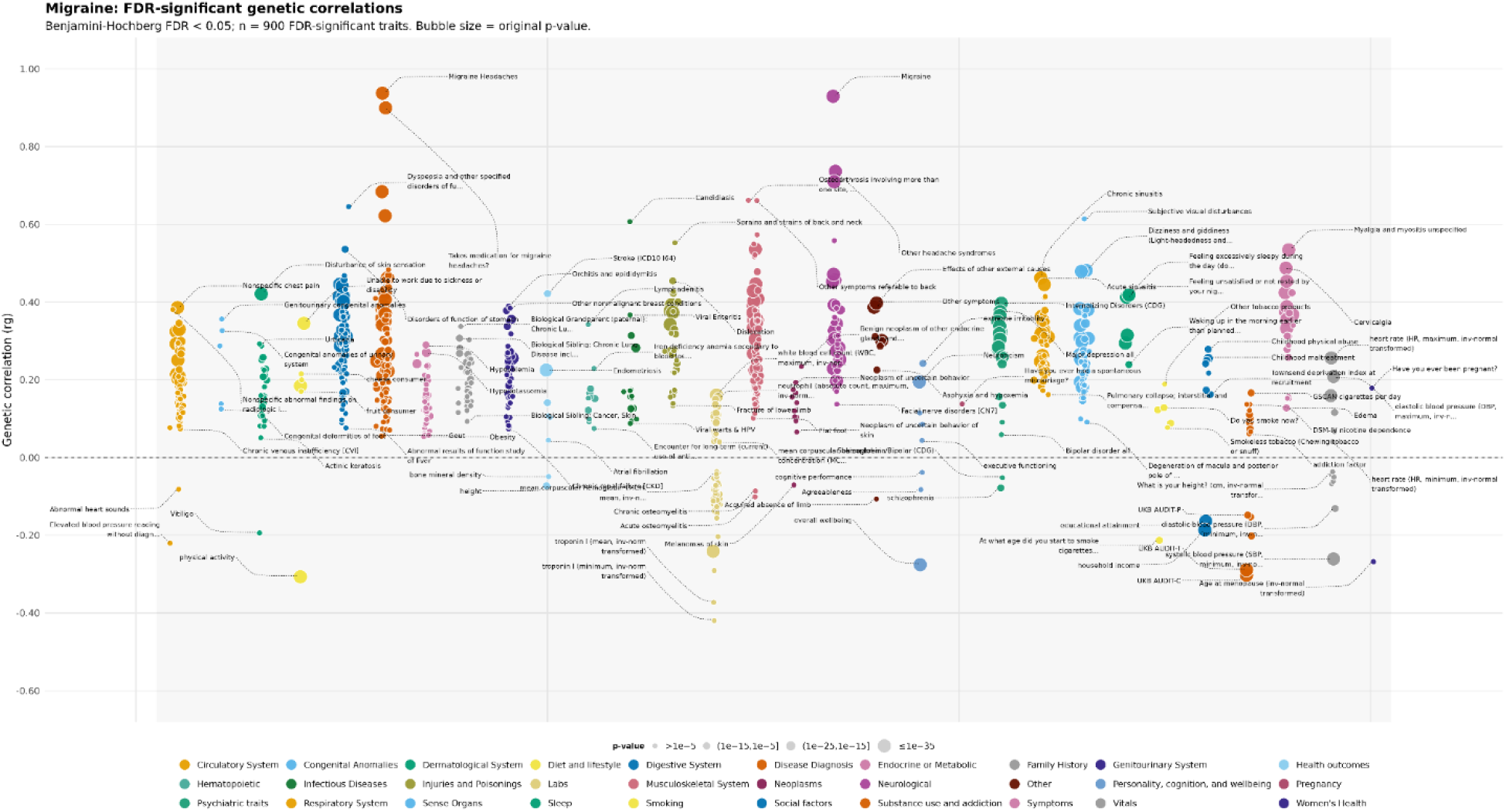
Genetic correlations between the European ancestry migraine meta-analysis and other traits. Points represent FDR-significant genetic correlations (FDR-adjusted *p* < 0.05); point size reflects the original *p*-value and colors indicate phenotype categories.

### Mendelian Randomization (MR)

Significant traits from the genetic correlation analyses were utilized downstream for MRlap. With migraine modeled as the exposure, 289 associations met the criteria of FDR-adjusted *p*<0.05 and no evidence of directional pleiotropy (MR-Egger intercept *p*≥0.05), including problematic alcohol use (UKB AUDIT-P; β=-0.101, SE=0.031, FDR-adjusted *p*=3.46×10^−3^), endometriosis (β=0.086, SE=0.026, FDR-adjusted *p*=3.45×10^−3^), opioid use disorder (β=0.114, SE=0.041, FDR-adjusted *p*=1.31×10^−2^), electrolyte imbalance (β=0.040, SE=0.014, FDR-adjusted *p*=1.25×10^−2^), and hypertension (β=0.067, SE=0.022, FDR-adjusted *p*=6.38×10^−3^; Figure 3, Supplementary Table 13).

**Figure 3.**
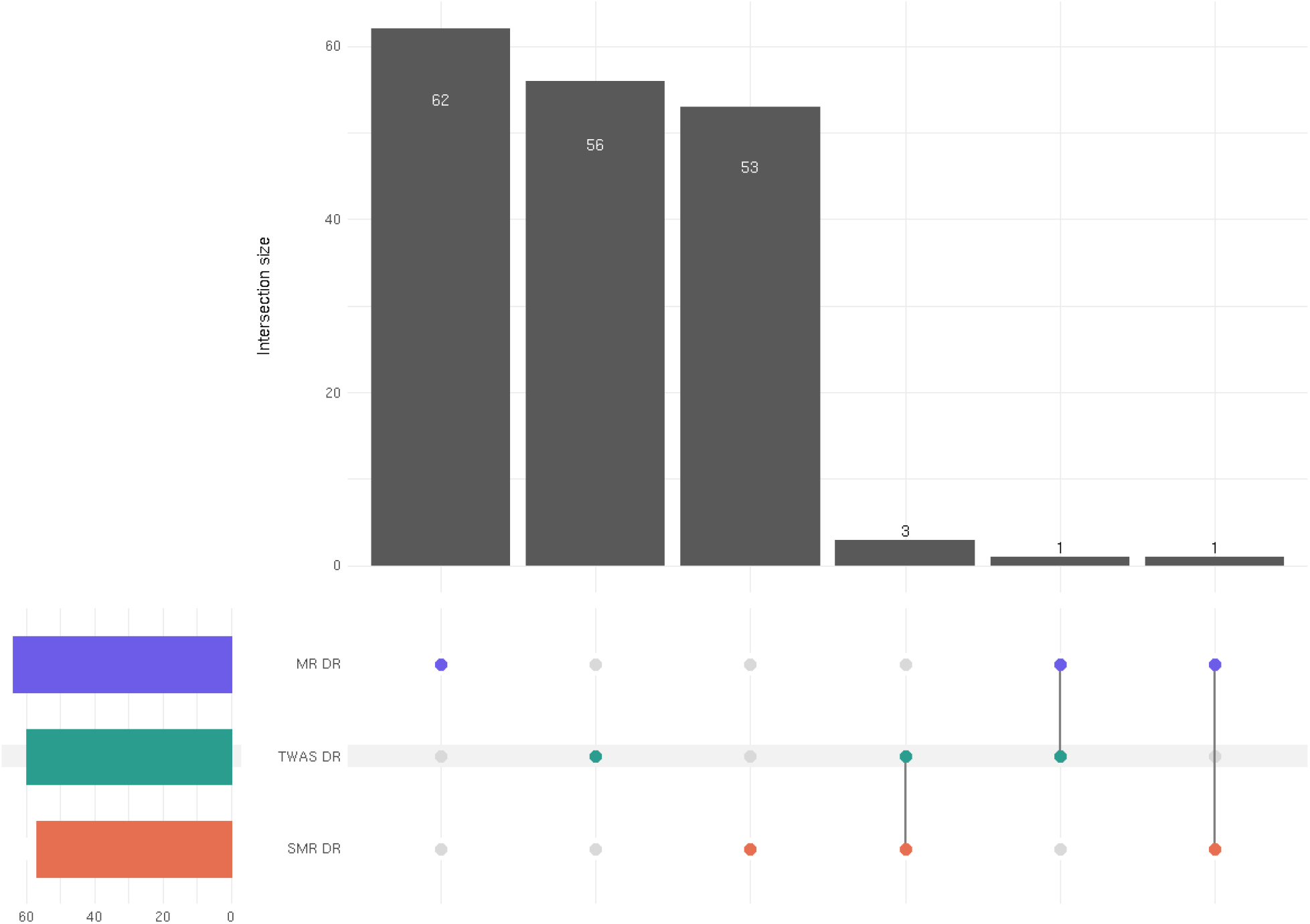
Comparison of drug-repurposing candidates across multiple analytic approaches. MR DR = Mendelian randomization drug repurposing results; TWAS DR = transcriptome-wide association study drug repurposing; SMR DR = summary-data-based Mendelian randomization drug repurposing results. Drug names were harmonized to parent ChEMBL v34 identifiers before overlap analysis; formulation variants mapping to the same parent compound were collapsed.

When considering migraine as the outcome, 101 traits met the criteria of FDR-adjusted *p*<0.05 and no evidence of directional pleiotropy (MR-Egger intercept *p*≥0.05), including vitamin D deficiency (β=0.044, SE=0.015, FDR-adjusted *p*=1.69×10^−2^), PTSD (β=0.197, SE=0.045, FDR-adjusted *p*=1.33×10^−4^), and esophagitis, GERD, and related diseases (β=0.179, SE=0.045, FDR-adjusted *p*=5.73×10^−4^). Full results are presented in Supplementary Table 14.

### Summary-data-based Mendelian Randomization (SMR) using pQTL Instruments

Using pQTL data, we performed SMR to identify proteins whose genetically regulated abundance showed evidence of association with migraine through shared causal or pleiotropic variants. SMR identified 23 genes with significant evidence of an association between migraine and genetically regulated protein abundance (Supplementary Table 15). These genes included *ITIH1*, *HSPA1A*, *CDH6*, *ABO*, *SEMA6C*, *RSPO3*, *TNC*, *VCAN*, *ATOX1*, *ARG1*, *TRIM26*, *EVI5*, *NRP2*, *KLB*, *NINJ1*, *NBL1*, *FABP3*, *PPCDC*, *ATXN2L*, *GSTM4*, *LIF*, *FUT3_FUT5*, and *TPPP3*.

### Transcriptome-wide Association Study (TWAS)

TWAS analyses in EUR across all tissues revealed 2,764 significant gene-trait associations (Supplementary Table 16), representing 541 unique genes (the same gene could be represented across multiple tissues). One hundred and eight genes identified in the TWAS overlapped with genes nearest to lead variants in the EUR migraine meta-analysis. We then examined the TWAS-derived gene set using g:Profiler^19^ and identified enrichment for the following GO terms: protein binding, nitrobenzene metabolic process, cytoplasm, nucleoplasm, nuclear speck, and cell projection (Extended Figure 9).

### Drug Repurposing (DR) Analyses

Results from the SMR-based approach (SMR DR; Supplementary Table 17), 59 drug-gene associations corresponding to 57 unique medications were identified. These included some established migraine prophylaxis medications such as β-blockers (e.g., propranolol). Medications associated with cardiovascular effects (e.g., simvastatin) and hormonal functioning (e.g., estradiol valerate) were also significant.

With the TWAS-based approach (TWAS DR; Supplementary Table 18) revealed 75 prioritized gene set-medication associations corresponding to 60 unique medications. Repeated medication entries represent distinct iLINCS perturbational signatures associated with the same compound. The finding with the smallest *p-*value implicated the steroid prednisolone (*p*=7.82×10^−4^). Several nonsteroidal anti-inflammatory (e.g., naproxen) and psychiatric medications (e.g., citalopram) were identified. Some medications with migraine/headache listed as a side effect were also significantly associated (e.g., sildenafil). Hormone-relevant (e.g., oxymetholone) and immunosuppressant medications (e.g., azathioprine) were also identified.

We further assessed the clinical relevance of the list of medication candidates derived from the TWAS and SMR-based DR analyses by comparing them to medications prescribed within 90 days of a migraine and/or headache diagnosis in the MSMHD cohort (MSMHD clinical relevance assessment; Supplementary Table 19). We identified 77 DR candidate drugs that were also prescribed within 90 days of a headache and/or migraine diagnosis in the MSMHD cohort, thereby providing clinical context for many of the drug candidates identified in the TWAS- and SMR-based DR analyses while also highlighting potential drug purposing candidates including simvastatin and mifepristone.

In a parallel effort using the MR-based approach (MR DR; Supplementary Table 20) we used the EUR meta-analysis results, *excluding* the 23andMe cohort, and applied a previously described pipeline on genome-wide targets^20^. When restricted to targets modulated by medications with regulatory approval, MR DR revealed 73 unique drug targets. Some frequently utilized migraine medications (CGRP monoclonal antibodies) that we identified included eptinezumab. Another medication identified in MR DR with potential relevance included dasatinib which is a tyrosine-kinase inhibitor that is not used clinically for migraine but has been mechanistically associated with pathways relevant to migraine, including MAPK/ERK signaling involved in CGRP release.

Finally, we examined unique and shared features across the drug-repurposing analytic approaches (Figure 3). Given variability in naming nomenclature, medications in each set of results (TWAS DR, SMR DR, MSMHD clinical relevance assessment, and MR DR) were mapped to their respective ChEMBL v34^21^ identifiers. Although no overlap was observed across all three DR methods, sildenafil, alcohol, and cisplatin were identified by both SMR DR and TWAS DR, cyclosporine by both SMR DR and MR DR, and methotrexate by both TWAS DR and MR DR. Sildenafil, cyclosporine, and methotrexate were additionally supported by significant results in the MSMHD clinical relevance assessment.

### Phenome-wide Association Study (PheWAS)

PheWAS results revealed 119 significant associations (Figure 4; Supplementary Table 21). As expected, top findings pertained to migraine (β=0.357, SE=0.018, *p*=4.28×10^−88^), migraine with aura (β=0.354, SE=0.028, *p*=1.45×10^−35^), and other headache syndromes (β=0.214, SE=0.018, *p*=3.41×10^−33^). Diseases of the esophagus (β=0.108, SE=0.013, *p*=4.28×10^−17^) and pain-related phenotypes (e.g., chronic pain syndrome [β=0.245, SE=0.035, *p*=2.25×10^−12^] and nonspecific chest pain [β=0.098, SE=0.013, *p*=8.64×10^−15^]) were also identified. Other top findings were consistent with common migraine symptoms (e.g., nausea and vomiting [β=0.115, SE=0.014, *p*=2.22×10^−15^] and dizziness [β=0.096, SE=0.014, *p*=9.47×10^−12^]). Multiple mental health traits were also identified, including posttraumatic stress disorder (β=0.218, SE=0.029, *p*=1.17×10^−13^), substance addiction and disorders (β=0.128, SE=0.022, *p*=5.11×10^−9^), opiates and related narcotics causing adverse effects in therapeutic use (β=0.219, SE=0.037, *p*=1.96×10^−9^), major depressive disorder (β=0.086, SE=0.015, *p*=1.66×10^−8^), and anxiety disorders (β=0.083, SE=0.014, *p*=5.74×10^−9^).

**Figure 4.**
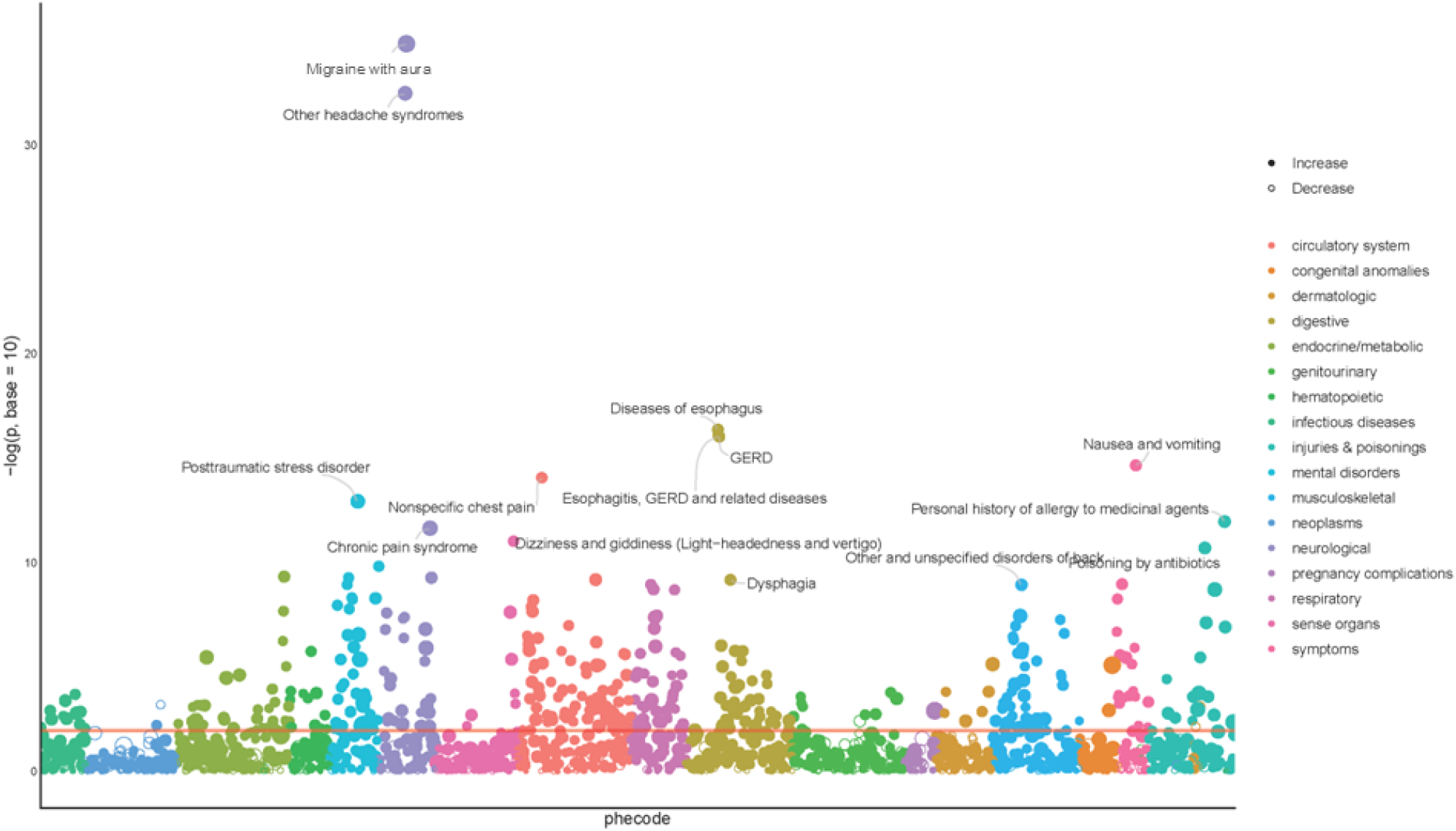
Phenome-wide association study of the European-ancestry migraine meta-analysis excluding 23andMe in the Mass General Brigham Biobank. Solid circles indicate positive associations and open circles indicate negative associations. Colors correspond to phecode categories.

### Epigenetic Age Acceleration (EAA)

Chronological age and Horvath DNAmAge descriptive information for migraine cases and controls in the MVP cohort are provided in Supplementary Table 22. In both ancestry groups, chronological age was significantly associated with Horvath DNAmAge (EUR: β=0.898, SE=0.002, *p*<2×10^-16^, *R*^2^_*adj*_; AFR: β=0.929, SE=0.003, *p*<2×10^-16^, *R*^2^_*adj*_). There were no significant associations between migraine case status and EAA in either EUR or AFR (Supplementary Table 23). Sex was a significant predictor of EAA, with female sex associated with decelerated aging in both EUR (β=-0.699, SE=0.114, *p*=9.88×10^-10^) and AFR (β=-0.408, SE=0.132, *p*=0.002).

## Discussion

The present study extends the genetic investigation of migraine by incorporating three novel datasets (AoU, MSMHD, and YP; 327,603 participants) together with recently performed migraine GWAS in additional cohorts (e.g., MVP) into the largest EUR, AFR, and multi-ancestry (EUR and AFR) migraine meta-analysis conducted to date, thereby increasing the EUR sample size to 2,300,068 participants and the total trans-ancestry sample size to ∼2.5 million. This analysis more than doubled the number of independent lead variants for EUR (311 vs. 123^4^), replicating many previously identified loci and identifying numerous novel associations. These differences should be interpreted in light of the more stringent LD-based pruning and locus-definition strategy used in the previous study^4^, which collapsed correlated signals using iterative *p*-value ranked pruning (*r²*>0.10) and defined loci using high-LD blocks (*r²*>0.60) compared to the FUMA default approach applied here. In AFR ancestry, two GWS variants were identified from a meta-analysis including MVP, YP, AoU, and MSMHD. The trans-ancestry meta-analysis revealed 316 lead variants across EUR and AFR cohorts. Across post-GWAS analyses, we observed consistent genetic associations between migraine and traits related to mental health, gastrointestinal disorders, and chronic pain, supporting and extending known biology.

Although there was variability in ascertainment across cohorts (e.g., ICD codes in MSMHD, self-report in 23andMe, inclusion of headache phecodes in the MVP algorithmic definition), genetic correlations were high (>0.867), supporting that meta-analysis was appropriate. Across the full EUR meta-analysis, we identified 311 lead variants, the nearest genes to which aligned with 108 genes identified in the TWAS results. Many of these genes implicate pathways consistent with our current biological understanding of migraine (sensory neuronal signaling [e.g., *RERE*, *TSPAN12*], vascular tone regulation [e.g., *LRP1*, *ABO*], glutamatergic neurotransmission [e.g., *CACNA1A*, *DCDC2*], and inflammatory processes [e.g., *STAT6*, *RNF213*]). Fine-mapping further refined putative causal variants and implicated genes, converging on *TRPM8*, *RNF213*, and *ENDOV* (present in both GWAS and fine mapping). Although *TRPM8* and *RNF213* have been consistently linked with migraine, *ENDOV* (which plays a role in DNA/RNA repair) has been less frequently discussed in relation to migraine. However, it has been shown to be associated with broadly defined headache in previous GWAS^63^ and fine-mapping, although the posterior inclusion probability was below the threshold as a high-confidence finding (PIP<0.01)^22^. One gene, *MRVI1* (*IRAG1)*, was associated consistently across the GWAS, fine-mapping, and TWAS analyses. This gene has a role in vascular tone and inflammatory modulation, biologically plausible in the context of migraine. Some genetic findings support links between *MRVI1* (*IRAG1*) and migraine (e.g., evidence of colocalization between migraine risk and *MRVI1* expression)^23,24^. This gene has also been linked to cardiovascular disorders, which is notable given the genetic correlation, PheWAS, and MR associations between migraine and cardiovascular traits (e.g., diastolic blood pressure; ischemic heart disease). MR analyses further supported this relationship between migraine and cardiovascular traits, with multiple cardiovascular traits associated including positive associations with pulmonary heart disease. *MRVI1 (IRAG1)* has been previously implicated in cardiovascular functioning (arterial stiffness in GWAS^25^ and smooth muscle dysfunction in *IRAG1* exon-12 knock-out mice^26^). Three other genes identified across GWAS, fine mapping, and TWAS analyses were *PHACTR1*, *CARF*, and *SLC24A3*. Both *PHACTR1* and *SLC24A3* are long-established migraine-relevant genes^12^; however, the evidence for *CARF* is less extensive (although identified in previous GWAS^27^, data included in the present analyses). *CARF* is associated with BDNF levels (associated with pain modulation, cortical excitability^28^, and depression^29^) and BDNF serum levels have well-documented associations with migraine^30^. Moreover, antidepressants which are occasionally utilized in migraine treatment (in lower doses than typically used to treat psychiatric disorders) impact BDNF levels (e.g., fluoxetine^31^); however, these medications are also known to worsen headache/migraine for some^32^, further highlighting the complexity of migraine treatment and the need for individualized treatment approaches. Lifestyle interventions (e.g., physical activity) which are frequently encouraged for migraine symptom management have also been shown to affect BDNF levels^33^. These multi-method convergent signals support the biological relevance of these genes to migraine. Additionally, MAGMA results showed enrichment in cervix and uterus. Given the context of hormone-related drug-repurposing hits, these results support that migraine biology might be viewed as neurovascular-endocrine, rather than simply neurovascular. The prevalence of migraine among women is double that among men^34^, so this interpretation may be relevant to the development of migraine therapies.

We used several approaches to contextualize migraine genetic risk relative to that of other traits. Our genetic correlation analyses, together with the MR and MGB PheWAS results, revealed a broad multimorbidity profile and complex clinical presentation of migraine. A common pattern of comorbidity included associations between migraine and psychiatric disorders, including depression, PTSD, ADHD, and anxiety, which have been reported in prior studies^35,22^. Research on the relationship between migraine and psychiatric traits has largely focused on internalizing disorders, a pattern we also observed in our genetic correlation, MR analyses (migraine as exposure), and PheWAS analyses. Our results also revealed significant associations with several substance-use-related phenotypes (e.g., opioid use disorder, but also therapeutic use of opioids). Although opioids have been prescribed in the past for migraine pain^36^, concerns regarding patterns of overprescribing, misuse and dependence, and research demonstrating poorer clinical outcomes have resulted in a shift away from the use of opioids to prioritizing migraine-specific and non-opioid treatments as first-line interventions^37^. Moreover, accumulating evidence indicates that opioid use can exacerbate migraine symptoms and contribute to pain sensitization across migraine and other pain–related conditions^38^, consistent with clinical recommendations against opioids in migraine treatment. We also identified substantial genetic overlap between migraine and other pain-related conditions, including multisite chronic pain, back pain, and spondylosis. These associations may reflect shared pain-related biology, such that the observed opioid genetic correlation may arise from traits that are highly genetically correlated with migraine (e.g., chronic pain conditions) rather than from migraine itself. Inverse associations were identified for alcohol use in the genetic correlation, MR analyses, and PheWAS analyses, with migraine genetic liability being inversely associated with alcohol-use phenotypes (e.g., current alcohol use). This pattern has been repeatedly documented^39,40^ and is often highlighted in migraine treatment discussions of lifestyle modifications (e.g., exercise, diet, sleep hygiene) that may improve migraine frequency/symptoms^41^. Many individuals with migraine abstain from alcohol even without clinical guidance, as alcohol is commonly perceived as a migraine trigger^42^.

Gastrointestinal, metabolic, and cardiovascular phenotypes also demonstrated strong associations with migraine. Conditions such as GERD, esophageal disease, and gastritis are frequently comorbid with migraine and may reflect shared mechanisms involving autonomic dysregulation, hypersensitivity, and gut-brain axis signaling^43^. Some gastrointestinal associations may partly reflect consequences of common migraine symptoms themselves (e.g., nausea and vomiting during migraine attacks may exacerbate or precipitate reflux). The associations with metabolic factors including obesity aligns with evidence implicating mechanisms involving insulin resistance, adipokine signaling, and low-grade systemic inflammation^44^. Cardiovascular features such as chest pain and ischemic heart disease were also identified, likely reflecting vascular and autonomic mechanisms that overlap with migraine^45^. Electrolyte imbalance may also impact cardiovascular physiology given the relationship between electrolytes and vascular tone.

Medication management for migraine is often characterized by a slow personalization process due to trial-and-error, clinical complexity, and shifting therapeutic needs as symptoms change over time. Given this complexity, we employed a comprehensive drug-repurposing analytic framework using multiple genetically informed approaches. Across methods, we observed convergent evidence again implicating inflammatory, vascular, hormonal, and neuropsychiatric pathways in migraine. TWAS- and SMR-based analyses highlighted anti-inflammatory medications, vascular modulators, and hormone-related compounds, alongside established migraine treatments such as CGRP-targeting therapies. The MR drug-repurposing approach further prioritized immune- and kinase-related targets, with several tyrosine kinase inhibitors (e.g., dasatinib) and methotrexate emerging across TWAS DR and MR DR analyses. Results from our clinical relevance cross-reference approach using EHR clinical prescribing data from the MSMHD cohort provided “real-world” clinical context for many drug candidates, as several genetically prioritized medications were also prescribed within 90 days of a migraine or headache diagnosis in that cohort. These findings reinforce pathways already implicated by existing migraine treatments (e.g., CGRP signaling, vascular tone, hormonal modulation), while also highlighting additional immune and kinase-related targets that may warrant future therapeutic investigation. Some identified compounds list migraine or headache as potential *adverse* effects (e.g., imatinib, sildenafil) indicating a possible good match for biology but a lack of therapeutic potential. Simvastatin and mifepristone (identified in the SMR DR analysis and observed in clinical prescribing in the MSMHD cohort) represent mechanistically plausible candidates; however, their signals may partly reflect co-occurring cardiovascular diagnoses or treatment of underlying symptoms. These patterns are also reflected across our PheWAS and MR analysis. Simvastatin influences endothelial function and vascular tone, and a randomized, placebo-controlled trial combining simvastatin with vitamin D reported reduced migraine frequency^46^. This result also aligns with our MR findings implicating vitamin D deficiency and several vascular-related traits when migraine was set as the outcome. Other statins, fluvastatin and atorvastatin, which have similar biological actions to simvastatin, were also identified in the TWAS-DR analyses. Mifepristone, a glucocorticoid-receptor antagonist, aligns with evidence implicating the HPA-axis and stress-related pathways in migraine. Mouse studies show that mifepristone may reduce cortical spreading depression (wave of neuronal and glial depolarization associated with migraine, particularly migraine with aura)^47^ and attenuate stress-induced migraine-like behaviors^48^. Finally, mifepristone’s actions on uterine and cervical tissues are notable considering our MAGMA results, which demonstrated significant enrichment in these tissues. These findings highlight potentially clinically relevant pathways, although substantial additional investigation is needed.

There was no significant association between migraine and epigenetic age acceleration (EAA) as measured by the Horvath epigenetic clock in either EUR or AFR. To our knowledge, this relationship (migraine and EAA) has not been studied previously. Research examining medication overuse headache (MOH) demonstrated significant decelerated aging using multiple methylation clocks^49^. This pattern may reflect immune-metabolic dysregulation which could dampen or blunt typical age-related methylation changes, thereby potentially reflecting a “younger” age compared to controls despite underlying biological alterations (e.g., chronic low-grade inflammation). This may also occur in migraine; more research is warranted. Comorbidity should be a consideration in future investigations of DNAm aging in migraine in addition to lifestyle factors (e.g., dietary variables, alcohol use) which have also been shown to strongly impact DNA methylation signatures^50^. MR results with migraine included as the outcome demonstrated a significant inverse association with obesity liability, which may reflect complex relationships between migraine, obesity-related biology, and behavioral or clinical factors associated with migraine management. Future research should incorporate these factors into DNAm aging analytic models.

Taken together, these findings align with extensive phenotypic and genetic evidence demonstrating that migraine is not solely a neurovascular disorder but associated with a broader array of medical (including endocrine-related) and psychiatric conditions. The convergence of gastrointestinal, cardiovascular, metabolic, inflammatory, and pain-related associations suggests that migraine may reflect a broader systemic biology, consistent with the heterogeneous symptom presentation observed clinically and the multifactorial nature of effective intervention strategies. Symptom patterns and treatment responsiveness change across the lifespan^51^ (e.g., a migraine medication may be effective when people are younger but they may require additional or different treatments with age). Moreover, given evidence that symptom trajectories evolve over time and the pronounced sex differences in migraine, particularly in relation to hormonal transitions such as pregnancy and menopause^34^, longitudinal examination and sex-stratified analyses are particularly warranted.

These findings should be interpreted in the context of several limitations. First, migraine phenotyping varied across cohorts, reflecting differences in ascertainment methods (e.g., ICD codes, self-report, and EHR-derived algorithms). In MVP, the phenotype was algorithmically defined and included headache-related phecodes. Although this approach may introduce some misclassification, the PheMED estimates and high genetic correlations suggest that any resulting bias is likely modest, and prior work has shown that many migraine cases are captured under headache codes. Other recent studies have taken a more restrictive approach by excluding other headache disorders to increase specificity^35^, underscoring the broader challenge of phenotype heterogeneity in migraine genetics. Additional phenotyping concerns include possible under- and misdiagnosis, as individuals who meet criteria for migraine may self–identify their symptoms as tension headache or other non-migraine headache conditions. Such misclassification can bias downstream analyses, reducing power and underscoring the need for nuanced and harmonized phenotyping in future work. Heterogeneity is also a concern within the migraine phenotype itself, as multiple migraine subtypes are subsumed under the migraine umbrella of migraine-relevant ICD-9/10 codes (e.g., hemiplegic migraine, migraine with/without cerebral infarction). There may be meaningful biological and genetic differences across migraine subtypes. Second, large sample sizes are useful for well-powered gene mapping studies even in the context of phenotypic heterogeneity, as reflected in the high genetic correlations across cohorts, but future work should explicitly disentangle phenotypic similarities and differences. Genetically dissecting distinctions between migraine with and without aura, chronic versus episodic migraine, and comorbid symptom profiles would be of great biological and clinical interest. Previous work identifying migraine subgroups based on comorbidity profiles has revealed meaningful differences across groups with both empirical and clinical implications, further underscoring the importance of phenotypic nuance in migraine^7^. Third, some degree of sample overlap across cohorts is likely, although the magnitude of this overlap cannot be determined. Our use of METACARPA^52^ in the meta-analytic framework was designed to mitigate this concern, by adjusting effect estimates and standard errors for unknown sample overlap, thereby preventing inflation of association statistics. Fourth, our analyses did not examine sex-specific genetic effects. Given the markedly higher prevalence of migraine among women and the well-documented role of hormonal pathways (including in this study), sex-stratified analyses represent an important next step. Similarly, efforts across additional ancestry groups are needed to provide better generalizability. Finally, the present study provides a broad overview of the genetic relationships between migraine and a wide range of traits but more work will be required to clarify causal mechanisms and directionality. For example, several traits demonstrated significant associations when migraine was set as both the exposure and the outcome in the MR analyses, highlighting complex interrelationships. These findings add to the mounting evidence suggesting systemic associations, pleiotropic relationships, and complex multimorbidity profiles between migraine and numerous psychiatric and medical traits; however, further investigation is critical to elucidate the imbricating nature of these variables.

In conclusion, results from this study substantially advance our understanding of the genetic architecture of migraine across EUR and AFR ancestry participants. By integrating newly generated GWAS with large existing resources, prioritizing candidate genes and variants through TWAS and fine-mapping analyses, and characterizing the broader multimorbidity landscape through genetic correlation and PheWAS analyses, this work provides a comprehensive framework for future biological investigation. As larger and more deeply phenotyped cohorts become available, particularly in underrepresented populations, future research should evaluate sex-, ancestry-, and subtype-specific effects and continue to advance genetically informed therapeutic discovery.

## Methods

### GWAS Analyses

#### All of Us Research Program (AoU)

Whole genome sequence data from AoU Version 8 was used. Briefly, we applied the following quality control (QC) thresholds: Variants with a minor allele frequency (MAF)<0.01 or Hardy-Weinberg Equilibrium (HWE) *p*<1×10^−6^ were removed. Covariates included age, sex, and 10 principal components (PCs) for EUR and 14 PCs for AFR ancestry.

Participants with Systematized Nomenclature of Medicine (SNOMED) code 37796009 in the EHR or those who selected ‘self’ to the ”Personal and Family Health History” survey question: “Including yourself, who in your family has had migraine headaches?”, were designated as cases. Controls included individuals with EHR data but no migraine SNOMED code and/or completed the ”personal and family health history” survey but did not respond with “self” to the self-report migraine item. The AoU cohort was predominately female (60.20%). Analyses were conducted separately for EUR (45,330 cases and 171,327 controls) and AFR (8,332 cases and 53,348 controls).

#### Mount Sinai Million Health Discoveries Program (MSMHD)

MSMHD (Mount Sinai Health System, New York, NY) has linked EHR and genotype data^1^. Participants with International Classification of Diseases (ICD) 9CM/10CM diagnoses of 346*, or G43* codes were designated as cases. MSMHD participants were genotyped on the Illumina Global Screening Array (GSA) and the multiplatform (combining GSA and custom automated workflow for genotyping-by-sequencing [GxS] developed at the Regeneron Corporation). Variants with MAF<0.01, HWE *p*<1×10^−6^, or poor imputation quality (MACH *R*^2^<0.3) were removed. Covariates included in the logistic GWAS included age, sex, and the first 10 PCs. The MSMHD cohort was predominately female (56.56%). Analyses were performed separately for EUR (1,301 cases and 20,764 controls) and AFR cohort (1,363 cases and 14,533 controls) participants.

#### Yale-Penn (YP)

YP participants were recruited at five sites across the Eastern United States for genetic studies of substance use disorders^2^. The participants were deeply phenotyped for Diagnostic and Statistical Manual of Mental Disorders, Fourth Edition (*DSM-IV-TR*^3^) defined psychiatric traits and medical and social history using the Semi-Structured Assessment for Drug Dependence and Alcoholism (SSADDA)^4^. Participants were asked questions regarding their medical history which included the following item: “Has a doctor ever told you that you have (had): Migraine headaches?”. A response of ‘Yes’ was considered a case. Three genotyping arrays were used across different enrollment periods: Illumina HumanOmni1-Quad v10 microarray, Illumina HumanCoreExome array, and Illumina Multi-Ethnic Global array. For QC, variants were removed if MAF<0.01, variant-level missingness >0.01, or HWE *p*<1×10^−6^. GWAS were performed using GEMMA univariate linear mixed modeling for each cohort (based on array [no sample overlap across arrays]) to account for the high degree of familial relatedness in the cohort^5^. Covariates included in the logistic GWAS were age, sex, and the first 10 PCs. The YP cohort was predominately male (54.46%). Analyses were performed separately for EUR (822 cases and 4,569 controls) and AFR (833 cases and 5,081 controls) participants. Results were then meta-analyzed within ancestry across the three YP cohorts using METAL^6^.

#### Overview of cohorts included in Meta-analysis

Five individual cohorts (AoU, 23andMe, MSMHD, Million Veteran Program [MVP], and YP) and seven cohorts included in a prior study^7^ (deCODE Genetics, Copenhagen Hospital Biobank [CHB] Study, Danish Blood Donor Study [DBDS], UK Biobank [UKB], FinnGen, Intermountain Healthcare [HerediGene], and HUSK) were included in the meta-analyses for EUR participants, a total of 206,893 migraine cases and 2,093,175 controls. Four datasets were included in the AFR ancestry meta-analysis (MVP, AoU, MSMHD, YP), a total of 22,115 cases and 178,626 controls. In accordance with 23andMe policy restricting data access to investigators listed on the 23andMe Statement of Work, this cohort was excluded from certain downstream analyses (phenome-wide association study and the Mendelian randomization-based drug-repurposing analyses). Migraine ascertainment across studies included electronic health records (EHR; defined by ICD and SNOMED codes), interview, and self-report survey responses (Table 1). Full details regarding each study and GWAS procedures (excluding datasets described in *GWAS Analyses* above), are included in the Supplementary Materials. Prior to meta-analysis, summary statistics were harmonized across cohorts using the MungeSumstats^8^ (R package) which addressed duplicates, ensured alleles were aligned across cohorts, applied MAF filtering (<0.01 for all cohorts) and converted summary statistics on Build 38 to Build 37 using UCSC Liftover^9^.

**Table 1.**
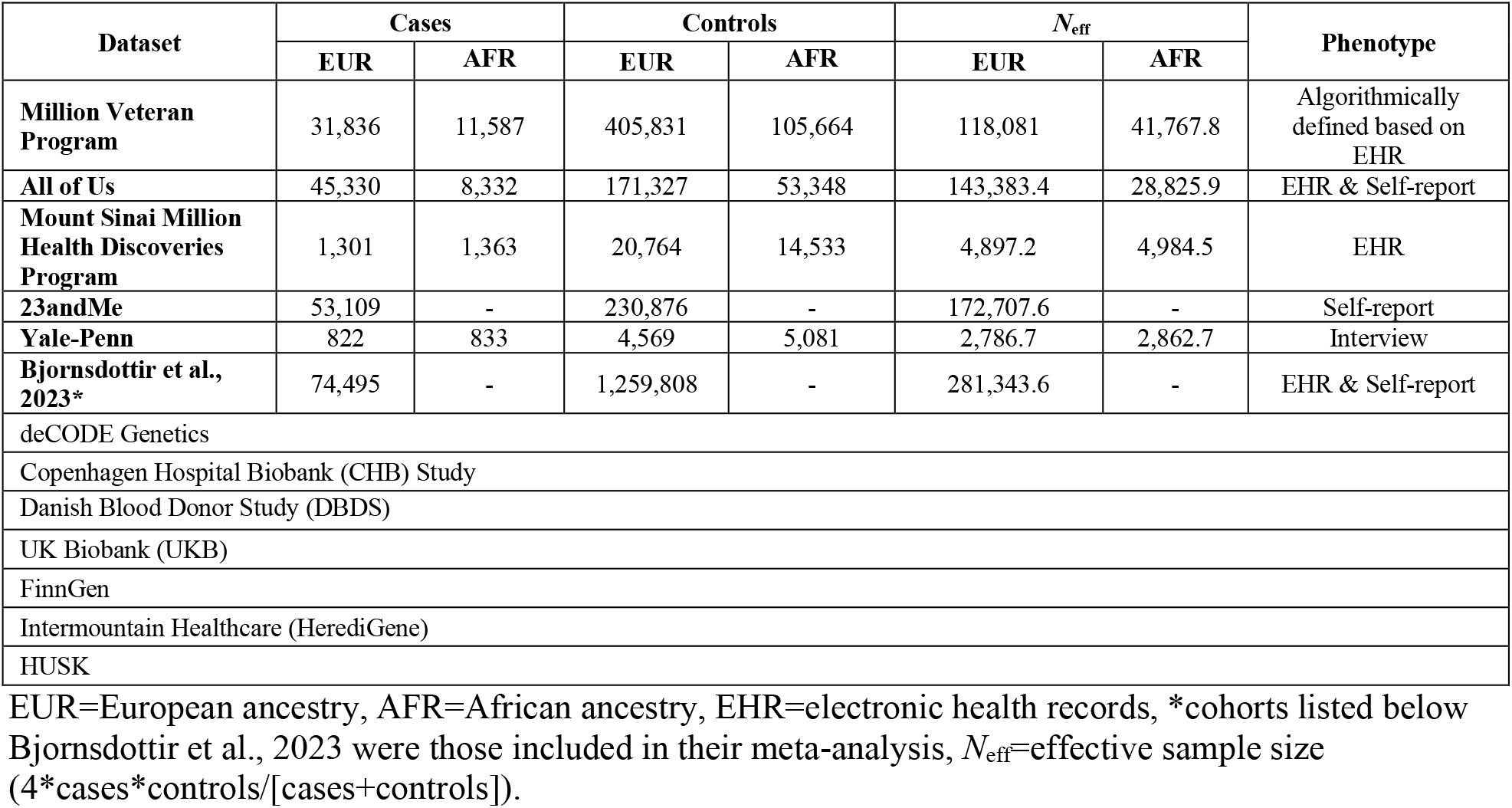
Sample sizes and phenotype ascertainment for cohorts included in the migraine meta-analysis.

#### Phenotypic Harmonization, SNP-based Heritability Estimates, and Genetic Correlations between Cohorts and across Ancestries

Migraine ascertainment varied across cohorts (e.g., self-report, EHR, interview) so Phenotypic Measurement of Effective Dilution (PheMED) was performed to assess potential bias associated with phenotypic dilution (noise associated with reduced specificity in case definitions)^10^. As recommended, we selected the cohort with the lowest expected likelihood of overlap to serve as the reference^7^ in the EUR PheMED analyses and individually compared each cohort to this reference to account for potential bias associated with sample overlap^10^. PheMED analyses were not performed for MSMHD or YP due to nonsignificant SNP-based heritability estimates identified in both EUR and AFR GWAS. For AFR, only MVP and AoU were included in the PheMED analyses with MVP serving as the reference.

We used linkage disequilibrium score (LDSC) regression to calculate liability-scale SNP-based heritability (*h²_SNP_*) estimates for each cohort (population prevalence 0.15^11^). For EUR, we used the EUR subset of 1KG Phase 3 LD reference panel^12^ and for AFR we used LD scores calculated from 10,000 random AFR individuals in AoU via cov-LDSC (to address systematic underestimation in LDSC for admixed populations^13^). LDSC was also used to calculate genetic correlations across cohorts to aid in determining whether meta-analysis would be appropriate (i.e., significant inter-cohort genetic correlations would indicate that each GWAS captures consistent genetic signals).

To examine trans-ancestry genetic correlations between EUR and AFR meta-analysis results, we utilized Popcorn^14^, which accounts for ancestry-specific differences in LD structure, allele frequencies, and sample sizes when analyzing GWAS summary statistics. We evaluated cross-ancestry similarity in genetic architecture using the genetic-impact correlation (PGI), which quantifies the correlation of effect sizes across ancestries weighted by ancestry-specific allele frequencies, and the genetic-effect correlation (PGE), which estimates the correlation of effect sizes without weighting, between EUR and AFR.

#### Meta-analysis

To account for case-control imbalance, effective sample sizes (*N*_eff_) for each cohort were used when performing the meta-analysis (Table 1). Due to potential unknown overlap across samples (individuals participating in multiple studies included in the meta-analyses [e.g., MVP, AoU, MSMHD, 23andMe, Intermountain Healthcare]), which could inflate association estimates, we utilized META-analysis in C++ Accounting for Relatedness, using arbitrary Precision Arithmetic (METACARPA^15^), which corrects for overlap by estimating the covariance structure between studies and incorporating it into the meta-analytic weights, producing overlap-adjusted effect sizes. The value provided in the “p_corrected” column of the results file was used as the GWAS *p*-value for each variant. The EUR analyses included the following cohorts: AoU, 23andMe, MSMHD, MVP, YP, and datasets from a previously published meta-analysis^7^. The AFR analyses included the following cohorts: AoU, MSMHD, MVP, and YP. METAL^6^ was used to perform the trans-ancestry (EUR and AFR) meta-analysis using the results for the EUR and AFR meta-analysis results from METACARPA^15^.

EUR migraine meta-analysis results including 23andMe were used in the post-GWAS *in silico* analyses except for the PheWAS and the Mendelian randomization-based drug-repurposing analyses which included all cohorts *except* 23andMe. The remaining analyses focused on the EUR sample (except for epigenetic age acceleration analyses which were completed in both EUR and AFR) due to the limited statistical power and few significant associations observed in the AFR meta–analysis.

#### Gene Mapping and Functional Annotation

Gene mapping and functional annotation of meta-analysis results were completed using the Functional Mapping and Annotation of Genome-Wide Association Studies (FUMA) platform (Version 1.5.4^16^). with a threshold of *r*^2^≥0.60 used to define independent significant SNPs and *r*^2^≥0.10 to define lead SNPs (default settings). A maximum distance of 10 kb was used for positional mapping of variants to genes. We also used the MAGMA analysis^17^ within the FUMA platform to assess tissue-specific expression enrichment. We also assessed enrichment of variants identified in the GWAS with previously reported associations in the GWAS Catalog^18^.

#### LD-aware SNP overlap analysis

Lead SNPs from the current meta-analysis were compared with variants reported in the most recent prior EUR ancestry migraine common variant meta-analysis^19^ incorporating local LD structure. Pairwise LD was calculated using the 1KG EUR reference panel^12^ using PLINK 1.9^20^ with a 1 Mb window and no minimum *r²* threshold (for complete LD coverage). LD pairs were retained only when at least one SNP originated from the present meta-analysis and at least one from the previous study. We then restricted LD pairs to those with *r²*≥0.40. SNPs from the current meta-analysis appearing in these LD pairs were classified as overlapping, whereas those without LD overlap at *r²*≥0.40 were designated putatively novel.

#### Fine Mapping

We performed fine mapping using echolocatoR (v2.0^21^), a suite of R packages that implements multi-method fine mapping. We utilized the echofinemap package to perform SNP-level fine mapping using multiple methods (SuSiE^22^, FINEMAP^23^). The EUR 1KG reference panel^12^ was used and we restricted to regions ±500 kb per genome-wide significant (GWS) SNP in our EUR migraine meta-analysis results. We prioritized consensus SNPs that were included in the 95% credible sets identified by both SuSiE and FINEMAP. High-confidence candidate variants were defined as those with a mean SNP-level posterior probability (PP) ≥0.95, calculated as the average of the SuSiE PP and FINEMAP SNP-level PP. Because ±500 kb windows per GWS SNP could overlap, the same variant could be identified in multiple locus-specific analyses. Therefore, we retained the fine mapping result with the highest mean SNP-level PP. SNPs were mapped to protein-coding genes (Ensembl GRCh37.87), while variants that could not be mapped to a protein-coding gene were assigned to the nearest transcription start site.

#### Genetic Correlations

We evaluated genetic correlations using the EUR migraine meta-analysis results; comparators were MVP traits included in a previously published GWAS-by-PheWAS (gwPheWAS)^24^, comprising 1,531 phenotypes across major clinical domains. As most psychiatric traits were not represented in the MVP gwPheWAS release, we also incorporated summary statistics for 91 traits that capture mental health and related outcomes of interest. We accounted for multiple correlated traits and measurements by applying FDR correction and considered those with an FDR-adjusted *p*<0.05 as significant.

#### Mendelian Randomization (MR)

FDR-significant traits from the genetic correlation analyses were further examined using Mendelian randomization with MRlap, an R package for two-sample MR that accounts for sample overlap^25^. We evaluated associations for migraine as both an exposure and an outcome, considering the following parameters: number of instruments, MRlap effect estimates and corresponding *p-*values, and MR-Egger intercept *p*-values. For each exposure-outcome pair, we report in *Results* the MRlap-corrected estimate when the test for difference between the corrected and uncorrected models resulted in *p*<0.05, indicating that the correction meaningfully altered the estimate. For difference tests with *p≥*0.05, we reported the observed (uncorrected) estimate. We applied an FDR correction to the *p*-values corresponding to the reported estimates (i.e., corrected vs. observed depending on the difference test *p*-value). We also flagged results with an MR-Egger intercept *p*<0.05 as showing evidence consistent with directional pleiotropy. Associations were considered significant without evidence consistent with directional pleiotropy when FDR-adjusted *p*<0.05 and the MR-Egger intercept *p≥*0.05.

#### Summary-data-based Mendelian Randomization (SMR) using pQTL Instruments

We performed SMR, which tests whether the association between a molecular trait (e.g., protein abundance) and a phenotype is consistent with a shared causal or pleiotropic variant, using the Complex Trait Genetics Virtual Lab (CTG-VL)^26^ platform (UK Biobank Plasma Proteomics Project). We then applied an FDR adjusted *p*<0.05 to identify significant associations. To distinguish putative causal signals from LD-driven associations, we applied the heterogeneity in dependent instruments (HEIDI) test, excluding associations with significant heterogeneity (*p*<0.05).

#### Transcriptome-wide Association Study (TWAS)

We used FUSION to perform the TWAS, integrating genetically predicted gene expression with summary statistics to determine gene-trait associations^27^. TWAS analyses were performed across all available tissues (*N*=49)^28^, with a Bonferroni-corrected significance threshold of 1.7×10^−7^ including all gene-tissue models (0.05/300,138 [number of gene-tissue pairs]). We then examined the TWAS-derived gene set using g:Profiler^29^ to identify enrichment of Gene Ontology (GO) terms.

#### Drug Repurposing (DR) Analyses

We utilized a multi-method approach to DR to reduce method-specific bias and identify convergence across methodologies. First, we used a TWAS–based approach (TWAS DR)^30^, using significant gene sets identified from our TWAS results. We ranked them by their TWAS z–scores, with the top 100 upregulated and top 100 downregulated genes selected to represent the disease–associated transcriptional signature. These gene-expression signatures were queried against the iLINCS drug-perturbagen database. Compounds were considered prioritized drug-repurposing candidates if *p*<0.05 and their perturbation signatures reversed the genetically regulated gene expression disease signature, defined as negative correlation coefficient or concordance value.

For the genetically regulated proteome targets identified using the SMR-based approach (SMR DR), we excluded those with significant HEIDI tests (*p*<0.05) to account for potential confounding and selected genes that were significant in our SMR analyses (FDR adjusted *p*<0.05 and HEIDI *p≥*0.05). We mapped these genes to drugs using the Drug-Gene Interaction Database (DGIdb^31^) to identify compounds targeting the implicated genes and restricted the final list to drugs with existing regulatory approval.

Although the primary purpose of these analyses was drug-repurposing, we also evaluated whether the DR-identified drugs were already used for migraine as a step to assess clinical relevance, since re-identification of known migraine medications increases confidence that the approach is identifying biologically relevant repurposing candidates. To do this, we examined whether drug candidates emerging from the TWAS and SMR DR analyses appeared in the MSMHD EHR records of medications prescribed within 90 days of a migraine and/or headache ICD diagnosis. Headache diagnoses were included given the high rates of migraine misdiagnosis in primary care and the clinical overlap between these conditions^32^.

We further implemented a Mendelian Randomization-based methodology for drug-repurposing analyses (MR DR) as described previously^33^. These analyses were performed using the EUR meta-analysis results *excluding* the 23andMe cohort. Genetic instruments were derived from five QTL resources spanning gene expression and protein abundance. For GTEx v8^34^, independent cis-eQTLs within ±1 Mb of each transcription start site were identified using up to five rounds of conditional analysis, retaining primary and conditional signals with *p*<5×10^−8^ and extracting effect estimates across all tissues. eQTLGen instruments were obtained from publicly available summary statistics. Protein QTLs were taken from three sources: deCODE SOMAscan v4 GWAS (4,907 aptamers in 35,000 EUR individuals)^35^; the Fenland study’s genome-proteome-wide association analysis (4,775 protein targets in 10,708 EUR individuals)^36^; and the Atherosclerosis Risk in Communities (ARIC) cis-pQTL study (4,657 plasma proteins in 7,213 European-American individuals)^37^. The significant cis-pQTLs (FDR<0.05) within ±500 kb of each gene were retained.

Two-sample MR was performed for 16,915 protein-coding genes across all phenotypes using harmonized effect estimates from each QTL source. Single-variant instruments were analyzed using the Wald ratio, and multi-variant instruments using inverse-variance weighted MR, with MR-Egger applied when ≥3 instruments were available. Gene-trait associations were considered significant at a Bonferroni-corrected threshold of *p*<3.76×10^−7^ (0.05/132,875 gene-trait tests). The number of tests is larger than the number of unique protein-coding genes because the same gene could be evaluated across multiple tissues, resulting in multiple gene-trait tests per gene. Next, associations were excluded if they demonstrated evidence of heterogeneity using Cochran’s Q (*p*<0.01), had a MR-Egger *p*-value (*p*<0.05), or showed MR-Egger heterogeneity (*p*<0.05). For single-variant instruments, sensitivity analyses were not applicable. For associations with two instruments, we evaluated Cochran’s Q, and for tests with ≥3 instruments we evaluated Cochran’s Q, the MR-Egger *p*-value, and MR-Egger heterogeneity per the aforementioned thresholds. Significant genes were mapped to drug information in ChEMBL v34^38^. We prioritized medications with regulatory approval that modulate targets identified in the MR DR analyses.

#### Phenome-wide Association Study (PheWAS) of Polygenic Risk Score (PRS) for Migraine

To characterize the broader clinical correlates of migraine genetic risk, we performed a PheWAS of migraine PRS in the Mass General Brigham (MGB) Biobank^39^. MGB participants were genotyped using the Illumina Global Screening Array, with genotype QC and principal component procedures documented publicly. Analyses were restricted to EUR ancestry because of insufficient sample sizes in AFR ancestry. Genotypes were imputed on the Michigan Imputation Server using the Haplotype Reference Consortium panel. Post-imputation filters excluded variants with INFO<0.80, MAF<0.01, HWE *p*<1×10^-10^, or missingness >0.02. After QC, the dataset included 30,701 unrelated EUR individuals. ICD 9/10 codes were mapped to phecodes, and individuals with ≥2 occurrences of a phecode were classified as cases, those with no occurrences as controls, and those with a single occurrence were excluded. A total of 1,706 phecodes with at least 20 cases were analyzed, and a Bonferroni threshold of *p*<2.93×10^-5^ was applied to identify significant associations.

Migraine (EUR meta-analysis *excluding* 23andMe) PRS were generated using PRS-CS^40^ and PLINK 1.9^20^ and standardized prior to PheWAS. Analyses were performed using logistic regression for each phecode, adjusting for age, sex, and the top 10 genetic PCs.

#### Epigenetic Age Acceleration

In the MVP cohort, DNA methylation age (DNAmAge) estimates were calculated by the MVP Core Team using the Horvath epigenetic clock method based on 353 CpG sites and trained on Illumina 27K and 450K arrays across multiple tissue types^41^ via the ”methylclock” package in R (v4.3.2). Analyses included all EUR (1,279 cases and 26,217 controls) and AFR (723 cases and 10,635 controls) participants with available migraine and epigenetic data. We regressed DNAmAge on chronological age and used the residuals as a measure of epigenetic age acceleration (EAA). We then included the following predictors in a linear regression model predicting EAA: migraine case status (algorithmically defined, as described in the supplementary materials), sex, and seven cell types (B cells, CD4+ T cells, CD8+ T cells, eosinophils, monocytes, neutrophils, and natural killer cells).

## Data Availability

Summary statistics from the meta-analyses excluding 23andMe data are available through Figshare. Complete GWAS summary statistics for the 23andMe data are accessible to qualified researchers through 23andMe under a data-access agreement designed to protect participant privacy. Additional details and application instructions are available at https://research.23andme.com/collaborate/#publication

Cassie Overstreet, Marco Galimberti, Kazi Tanvir Hasan, Sarah Beck, Brian Ferolito, Murray B. Stein, Alexandre C. Pereira, Joseph D. Deak, J. Michael Gaziano, Daniel F. Levey, & Joel Gelernter. VA Million Veteran Program Core Acknowledgements for Publications provided in the Supplementary Materials.

## Acknowledgements

This research is based in part on data from the Million Veteran Program, Office of Research and Development, Veterans Health Administration and was supported by MP000, MVP092 and MVP069 as well as award #2IO1BX006482 and #5IK2BX005058. More details regarding this consortium are provided in the Supplementary Information. This publication does not represent the views of the Department of Veterans Affairs or the United States Government. We thank the participants of the Million Veteran Program for their contributions to this research.

We thank the research participants and employees of 23andMe Research Institute for their contribution to this study. Summary statistics for the 23andMe cohort were provided under a data-sharing agreement with 23andMe Research Institute.

We gratefully acknowledge the contributions of the participants in the All of Us Research Program.

We gratefully acknowledge the Mount Sinai Million Health Discoveries Program at the Icahn School of Medicine at Mount Sinai and its research participants for their invaluable contributions to this research.

We acknowledge the use of data from the database of Genotypes and Phenotypes (dbGaP; accession phs002453.v1.p1).

We thank the participants and research staff of the Yale-Penn study for their contributions.

## Author Contributions

C.O., G.A.P., D.F.L., and J.G. conceptualized and designed the study. C.O., G.A.P., M.G., K.T.H., S.B., J.H., S.S., B.F., Y. Zhou, Y. Zhang, E.I.W., J.D.D., A.C.P., and K.C. performed data analyses. C.O. prepared the manuscript. The VA Million Veteran Program contributed data and resources and supported data generation and cohort-level analyses. All authors contributed to interpretation of results and reviewed and approved the manuscript.

## Competing Interests

Dr. Stein has in the past 3 years received consulting income from Abbvie, ataiBeckley, Ananda Scientific, BigHealth, Biogen, Bionomics/Neuphoria, Boehringer Ingelheim, EmpowerPharm, Engrail Therapeutics, Jazz Pharmaceuticals, Karuna Therapeutics, Lundbeck, Lykos Therapeutics, Newleos Therapeutics, Orion Pharma, Otsuka US, PureTech Health, Roche/Genentech, Sage Therapeutics, Seaport Therapeutics, Sensorium Therapeutics, and Transcend Therapeutics. Stein has stock options in EpiVario, Newleos Therapeutics, and Oxeia Biopharmaceuticals. He has been paid for his editorial work on *Biological Psychiatry* (Deputy Editor), and *UpToDate* (Co-Editor-in-Chief for Psychiatry). He has also received research support from the National Institutes of Health, the Department of Veterans Affairs, and the Department of Defense. He is on the scientific advisory board of the Brain and Behavior Research Foundation and the Anxiety and Depression Association of America. Dr. Gelernter is paid for editorial work by the journal *Complex Psychiatry* (Karger). Dr. Kranzler is a member of advisory boards for Altimmune, Clearmind Medicine, and Niuvera Bio; a consultant to Sobrera Pharmaceuticals, Altimmune, Lilly, Ribocure, and Boehringer Ingelheim; and the recipient of research funding and medication supplies for an investigator-initiated study from Alkermes and company-initiated studies by Altimmune and Lilly. All other authors report no potential conflicts of interest.

**Extended Data Fig. 1 Manhattan plot of the European ancestry migraine meta-analysis *excluding* 23andMe.** The red dashed line indicates the genome-wide significance threshold (*p* = 5×10^-8^).

**Extended Data Fig. 2 Manhattan plot of migraine genome-wide association results in European ancestry participants from the All of Us Research Program (45,330 cases; 171,327 controls).** The red dashed line indicates the genome-wide significance threshold (*p* = 5×10^-8^).

**Extended Data Fig. 3 Manhattan plot of migraine genome-wide association results in African ancestry participants from the All of Us Research Program (8,332 cases; 53,348 controls).** The red dashed line indicates the genome-wide significance threshold (*p* = 5×10^-8^).

**Extended Data Fig. 4 Manhattan plot of migraine genome-wide association results in European ancestry participants from the Yale-Penn cohort (822 cases; 4,569 controls).** The red dashed line indicates the genome-wide significance threshold (*p* = 5×10^-8^).

**Extended Data Fig. 5 Manhattan plot of migraine genome-wide association results in African ancestry participants from the Yale-Penn cohort (833 cases; 5,081 controls).** The red dashed line indicates the genome-wide significance threshold (*p* = 5×10^-8^).

**Extended Data Fig. 6 Manhattan plot of the African ancestry migraine meta-analysis.** The red dashed line indicates the genome-wide significance threshold (*p* = 5×10^-8^).

**Extended Data Fig. 7 Manhattan plot of the trans-ancestry migraine meta-analysis.** The red dashed line indicates the genome-wide significance threshold (*p* = 5×10^-8^).

**Extended Data Fig. 8 Tissue-specific gene-expression enrichment for the European ancestry migraine meta-analysis using FUMA GENE2FUNC.** Significantly enriched DEG sets (Bonferroni-corrected *p* < 0.05) are highlighted in red.

**Extended Data Fig. 9 g:Profiler enrichment results for the TWAS-derived gene set.**

## References

1. Steiner, T. et al. Migraine remains second among the world’s causes of disability, and first among young women: findings from GBD2019. The journal of headache and pain 21, 137 (2020).

2. Eigenbrodt, A.K. et al. Diagnosis and management of migraine in ten steps. Nature Reviews Neurology 17, 501–514 (2021).

3. Trajanoska, K. et al. From target discovery to clinical drug development with human genetics. Nature 620, 737–745 (2023).

4. Hautakangas, H. et al. Genome-wide analysis of 102,084 migraine cases identifies 123 risk loci and subtype-specific risk alleles. Nature genetics 54, 152–160 (2022).

5. Steiner, T.J. & Stovner, L.J. Global epidemiology of migraine and its implications for public health and health policy. Nature Reviews Neurology 19, 109–117 (2023).

6. Headache Classification Committee of the International Headache Society (IHS) The International Classification of Headache Disorders, 3rd edition. Cephalalgia 38, 1–211 (2018).

7. Lipton, R.B. et al. Identifying natural subgroups of migraine based on comorbidity and concomitant condition profiles: results of the chronic migraine epidemiology and outcomes (CaMEO) study. Headache: The Journal of Head and Face Pain 58, 933–947 (2018).

8. Dresler, T. et al. Understanding the nature of psychiatric comorbidity in migraine: a systematic review focused on interactions and treatment implications. The journal of headache and pain 20, 51 (2019).

9. Society, A.H. The American Headache Society position statement on integrating new migraine treatments into clinical practice. Headache: The Journal of Head and Face Pain 59, 1–18 (2019).

10. Silberstein, S.D. et al. Pharmacological approaches to managing migraine and associated comorbidities—clinical considerations for monotherapy versus polytherapy. Headache: The Journal of Head and Face Pain 47, 585–599 (2007).

11. Grangeon, L. et al. Genetics of migraine: where are we now? The journal of headache and pain 24, 12 (2023).

12. Sutherland, H.G., Albury, C.L. & Griffiths, L.R. Advances in genetics of migraine. The journal of headache and pain 20, 72 (2019).

13. Bjornsdottir, G. et al. Rare variants with large effects provide functional insights into the pathology of migraine subtypes, with and without aura. Nature Genetics 55, 1843–1853 (2023).

14. Investigators, A.o.U.R.P. The “All of Us” research program. New England Journal of Medicine 381, 668–676 (2019).

15. Wojcik, G.L. et al. Genetic analyses of diverse populations improves discovery for complex traits. Nature 570, 514–518 (2019).

16. Gelernter, J. et al. Genome-wide association study of opioid dependence: multiple associations mapped to calcium and potassium pathways. Biological psychiatry 76, 66–74 (2014).

17. Horvath, S. DNA methylation age of human tissues and cell types. Genome biology 14, 3156 (2013).

18. Consortium, G.P. A global reference for human genetic variation. Nature 526, 68 (2015).

19. Raudvere, U. et al. g: Profiler: a web server for functional enrichment analysis and conversions of gene lists (2019 update). Nucleic acids research 47, W191–W198 (2019).

20. Ferolito, B.R. et al. Leveraging large-scale biobanks for therapeutic target discovery. Human Genetics and Genomics Advances 7(2026).

21. Mendez, D. et al. ChEMBL: towards direct deposition of bioassay data. Nucleic acids research 47, D930–D940 (2019).

22. Hautakangas, H. et al. Fine-mapping a genome-wide meta-analysis of 98,374 migraine cases identifies 181 sets of candidate causal variants. Nature Communications 17, 355 (2026).

23. Prüschenk, S., Majer, M. & Schlossmann, J. Novel functional features of cGMP substrate proteins IRAG1 and IRAG2. International Journal of Molecular Sciences 24, 9837 (2023).

24. Lou, J., Tu, M., Xu, M., Cao, Z. & Song, W. Plasma pQTL and brain eQTL integration identifies PNKP as a therapeutic target and reveals mechanistic insights into migraine pathophysiology. The Journal of Headache and Pain 25, 202 (2024).

25. Fung, K. et al. Genome-wide association study identifies loci for arterial stiffness index in 127,121 UK Biobank participants. Scientific reports 9, 9143 (2019).

26. Geiselhöringer, A. et al. IRAG is essential for relaxation of receptor-triggered smooth muscle contraction by cGMP kinase. The EMBO journal 23, 4222 (2004).

27. Gormley, P. et al. Meta-analysis of 375,000 individuals identifies 38 susceptibility loci for migraine. Nature genetics 48, 856–866 (2016).

28. Ren, K. & Dubner, R. Pain facilitation and activity-dependent plasticity in pain modulatory circuitry: role of BDNF-TrkB signaling and NMDA receptors. Molecular neurobiology 35, 224–235 (2007).

29. Cavaleri, D. et al. The role of BDNF in major depressive disorder, related clinical features, and antidepressant treatment: Insight from meta-analyses. Neuroscience & Biobehavioral Reviews 149, 105159 (2023).

30. Martins, L., Teixeira, A. & Domingues, R. Neurotrophins and migraine. Vitamins and hormones 104, 459–473 (2017).

31. Duman, R.S., Deyama, S. & Fogaça, M.V. Role of BDNF in the pathophysiology and treatment of depression: Activity-dependent effects distinguish rapid-acting antidepressants. European Journal of Neuroscience 53, 126–139 (2021).

32. Telang, S., Walton, C., Olten, B. & Bloch, M.H. Meta-analysis: second generation antidepressants and headache. Journal of affective disorders 236, 60–68 (2018).

33. Huang, T., Larsen, K.T., Ried-Larsen, M., Møller, N.C. & Andersen, L.B. The effects of physical activity and exercise on brain-derived neurotrophic factor in healthy humans: A review. Scandinavian journal of medicine & science in sports 24, 1–10 (2014).

34. Vetvik, K.G. & MacGregor, E.A. Sex differences in the epidemiology, clinical features, and pathophysiology of migraine. The Lancet Neurology 16, 76–87 (2017).

35. Gasperi, M. et al. A multi-ancestry meta genome-wide association study of migraine among veterans: associations with traumatic brain injury, depression, and post-traumatic stress disorder. Molecular psychiatry, 1–15 (2025).

36. Lipton, R.B. et al. Burden of increasing opioid use in the treatment of migraine: Results from the Migraine in America Symptoms and Treatment Study. Headache: The Journal of Head and Face Pain 61, 103–116 (2021).

37. Ashina, S. et al. Opioid Use among People with Migraine: Results of the OVERCOME (US) Study. Pain and Therapy 14, 1745–1763 (2025).

38. Johnson, J.L., Hutchinson, M.R., Williams, D.B. & Rolan, P. Medication-overuse headache and opioid-induced hyperalgesia: a review of mechanisms, a neuroimmune hypothesis and a novel approach to treatment. Cephalalgia 33, 52–64 (2013).

39. Błaszczyk, B. et al. Relationship between alcohol and primary headaches: a systematic review and meta-analysis. The Journal of Headache and Pain 24, 116 (2023).

40. Zhou, H. et al. Multi-ancestry study of the genetics of problematic alcohol use in over 1 million individuals. Nature Medicine 29, 3184–3192 (2023).

41. Nigade, A., Pathak, G., Baidya, M. & Bhatt, S. Lifestyle Modifications for the Management of Migraine Pain. in Management of Migraine Pain: Emerging Opportunities and Challenges 45–61 (Springer, 2024).

42. Onderwater, G., Van Oosterhout, W., Schoonman, G., Ferrari, M. & Terwindt, G. Alcoholic beverages as trigger factor and the effect on alcohol consumption behavior in patients with migraine. European Journal of Neurology 26, 588–595 (2019).

43. Chu, B.K., Karhu, E., Li, B. & Sonu, I. Gastrointestinal Symptoms and Comorbid Conditions in Migraine. Comorbid Conditions in the Treatment of Headache, 125–145 (2024).

44. Rivera-Mancilla, E., Al-Hassany, L., Villalón, C.M. & MaassenVanDenBrink, A. Metabolic aspects of migraine: association with obesity and diabetes mellitus. Frontiers in neurology 12, 686398 (2021).

45. Kurth, T. & Rist, P.M. Migraines and cardiovascular disease: mechanisms and methodological challenges. Nature Reviews Cardiology 20, 791–792 (2023).

46. Buettner, C. et al. Simvastatin and vitamin D for migraine prevention: a randomized, controlled trial. Annals of neurology 78, 970–981 (2015).

47. Shyti, R. et al. Stress hormone corticosterone enhances susceptibility to cortical spreading depression in familial hemiplegic migraine type 1 mutant mice. Experimental neurology 263, 214–220 (2015).

48. Hu, Y.-Y. et al. Glucocorticoid signaling mediates stress-induced migraine-like behaviors in a preclinical mouse model. Cephalalgia 44, 03331024241277941 (2024).

49. Kwiatkowska, K.M. et al. Whole blood DNA methylation signature of epigenetic aging in medication overuse headache. The Journal of Headache and Pain (2026).

50. Galkin, F., Kovalchuk, O., Koldasbayeva, D., Zhavoronkov, A. & Bischof, E. Stress, diet, exercise: common environmental factors and their impact on epigenetic age. Ageing Research Reviews 88, 101956 (2023).

51. Bigal, M.E. & Lipton, R.B. Migraine chronification. Current neurology and neuroscience reports 11, 139–148 (2011).

52. Southam, L. et al. Whole genome sequencing and imputation in isolated populations identify genetic associations with medically-relevant complex traits. Nature communications 8, 15606 (2017).

## Methods References

1. Wojcik, G.L. et al. Genetic analyses of diverse populations improves discovery for complex traits. Nature 570, 514–518 (2019).

2. Gelernter, J. et al. Genome-wide association study of opioid dependence: multiple associations mapped to calcium and potassium pathways. Biological psychiatry 76, 66–74 (2014).

3. American Psychiatric Association. *Diagnostic and statistical manual of mental disorders, text revision (DSM-IV-TR)*, (American Psychiatric Association, Washington, DC, 2000).

4. Pierucci-Lagha, A. et al. Diagnostic reliability of the Semi-structured Assessment for Drug Dependence and Alcoholism (SSADDA). Drug and alcohol dependence 80, 303–312 (2005).

5. Zhou, X. & Stephens, M. Genome-wide efficient mixed-model analysis for association studies. Nature genetics 44, 821–824 (2012).

6. Willer, C.J., Li, Y. & Abecasis, G.R. METAL: fast and efficient meta-analysis of genomewide association scans. Bioinformatics 26, 2190–2191 (2010).

7. Bjornsdottir, G. et al. Rare variants with large effects provide functional insights into the pathology of migraine subtypes, with and without aura. Nature Genetics 55, 1843–1853 (2023).

8. Murphy, A.E., Schilder, B.M. & Skene, N.G. MungeSumstats: a Bioconductor package for the standardization and quality control of many GWAS summary statistics. Bioinformatics 37, 4593–4596 (2021).

9. Hinrichs, A.S. et al. The UCSC genome browser database: update 2006. Nucleic acids research 34, D590–D598 (2006).

10. Burstein, D. et al. Detecting and adjusting for hidden biases due to phenotype misclassification in genome-wide association studies. *medrxiv* (2023).

11. Steiner, T.J. & Stovner, L.J. Global epidemiology of migraine and its implications for public health and health policy. Nature Reviews Neurology 19, 109–117 (2023).

12. Consortium, G.P. A global reference for human genetic variation. Nature 526, 68 (2015).

13. Luo, Y. et al. Estimating heritability and its enrichment in tissue-specific gene sets in admixed populations. Human molecular genetics 30, 1521–1534 (2021).

14. Brown, B.C., Ye, C.J., Price, A.L. & Zaitlen, N. Transethnic genetic-correlation estimates from summary statistics. The American Journal of Human Genetics 99, 76–88 (2016).

15. Southam, L. et al. Whole genome sequencing and imputation in isolated populations identify genetic associations with medically-relevant complex traits. Nature communications 8, 15606 (2017).

16. Watanabe, K., Taskesen, E., Van Bochoven, A. & Posthuma, D. Functional mapping and annotation of genetic associations with FUMA. Nature communications 8, 1826 (2017).

17. De Leeuw, C.A., Mooij, J.M., Heskes, T. & Posthuma, D. MAGMA: generalized gene-set analysis of GWAS data. PLoS computational biology 11, e1004219 (2015).

18. Buniello, A. et al. The NHGRI-EBI GWAS Catalog of published genome-wide association studies, targeted arrays and summary statistics 2019. Nucleic acids research 47, D1005–D1012 (2019).

19. Hautakangas, H. et al. Genome-wide analysis of 102,084 migraine cases identifies 123 risk loci and subtype-specific risk alleles. Nature genetics 54, 152–160 (2022).

20. Chang, C.C. et al. Second-generation PLINK: rising to the challenge of larger and richer datasets. Gigascience 4, s13742-015-0047-8 (2015).

21. Schilder, B.M., Humphrey, J. & Raj, T. echolocatoR: an automated end-to-end statistical and functional genomic fine-mapping pipeline. Bioinformatics 38, 536–539 (2022).

22. Zou, Y., Carbonetto, P., Wang, G. & Stephens, M. Fine-mapping from summary data with the “Sum of Single Effects” model. PLoS genetics 18, e1010299 (2022).

23. Benner, C. et al. FINEMAP: efficient variable selection using summary data from genome-wide association studies. Bioinformatics 32, 1493–1501 (2016).

24. Verma, A. et al. Diversity and scale: Genetic architecture of 2068 traits in the VA Million Veteran Program. Science 385, eadj1182 (2024).

25. Mounier, N. & Kutalik, Z. Bias correction for inverse variance weighting Mendelian randomization. Genetic Epidemiology 47, 314–331 (2023).

26. Cuellar-Partida, G. et al. Complex-Traits Genetics Virtual Lab: A community-driven web platform for post-GWAS analyses. *BioRxiv*, 518027 (2019).

27. Gusev, A. et al. Integrative approaches for large-scale transcriptome-wide association studies. Nature genetics 48, 245–252 (2016).

28. Pividori, M. et al. PhenomeXcan: Mapping the genome to the phenome through the transcriptome. Science advances 6, eaba2083 (2020).

29. Raudvere, U. et al. g: Profiler: a web server for functional enrichment analysis and conversions of gene lists (2019 update). Nucleic acids research 47, W191–W198 (2019).

30. Wu, P. et al. Integrating gene expression and clinical data to identify drug repurposing candidates for hyperlipidemia and hypertension. Nature Communications 13, 46 (2022).

31. Cannon, M. et al. DGIdb 5.0: rebuilding the drug–gene interaction database for precision medicine and drug discovery platforms. Nucleic acids research 52, D1227–D1235 (2024).

32. Lipton, R.B., Diamond, S., Reed, M., Diamond, M.L. & Stewart, W.F. Migraine diagnosis and treatment: results from the American Migraine Study II. Headache: The Journal of Head and Face Pain 41, 638–645 (2001).

33. Ferolito, B.R. et al. Leveraging large-scale biobanks for therapeutic target discovery. Human Genetics and Genomics Advances 7(2026).

34. Consortium, G. The GTEx Consortium atlas of genetic regulatory effects across human tissues. Science 369, 1318–1330 (2020).

35. Ferkingstad, E. et al. Large-scale integration of the plasma proteome with genetics and disease. Nature genetics 53, 1712–1721 (2021).

36. Pietzner, M. et al. Synergistic insights into human health from aptamer-and antibody-based proteomic profiling. Nature communications 12, 6822 (2021).

37. Zhang, J. et al. Plasma proteome analyses in individuals of European and African ancestry identify cis-pQTLs and models for proteome-wide association studies. Nature genetics 54, 593–602 (2022).

38. Mendez, D. et al. ChEMBL: towards direct deposition of bioassay data. Nucleic acids research 47, D930–D940 (2019).

39. Boutin, N.T. et al. The evolution of a large biobank at Mass General Brigham. Journal of Personalized Medicine 12, 1323 (2022).

40. Ge, T., Chen, C.-Y., Ni, Y., Feng, Y.-C.A. & Smoller, J.W. Polygenic prediction via Bayesian regression and continuous shrinkage priors. Nature communications 10, 1776 (2019).

41. Horvath, S. DNA methylation age of human tissues and cell types. Genome biology 14, 3156 (2013).

