## Extended Figures for "Genomic Architecture of Migraine: A Multi-ancestry GWAS Meta-analysis of 2.5 Million Participants"

**Extended Data Figures**

**Extended Data Fig. 1 | Manhattan plot of the European ancestry migraine meta-analysis *excluding* 23andMe.**

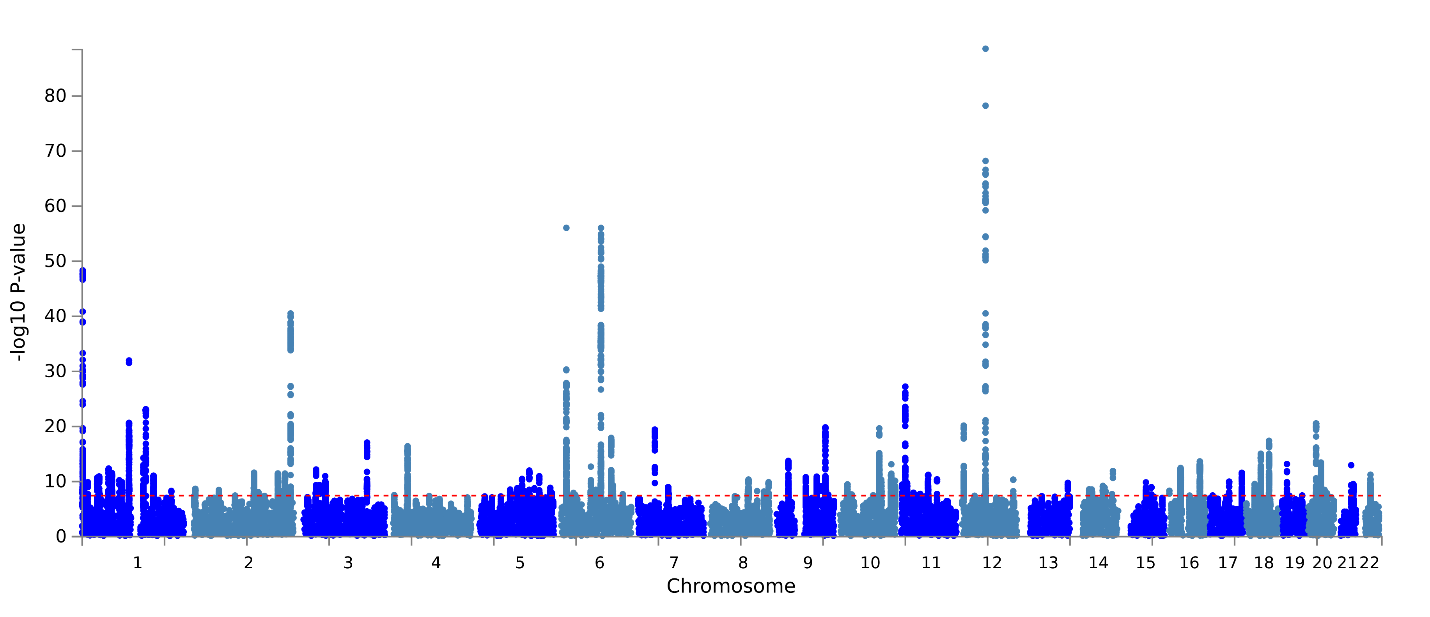

The red dashed line indicates the genome-wide significance threshold (*p* = 5×10⁻⁸).

**Extended Data Fig. 2 | Manhattan plot of migraine genome-wide association results in European ancestry participants from the All of Us Research Program (45,330 cases; 171,327 controls).**
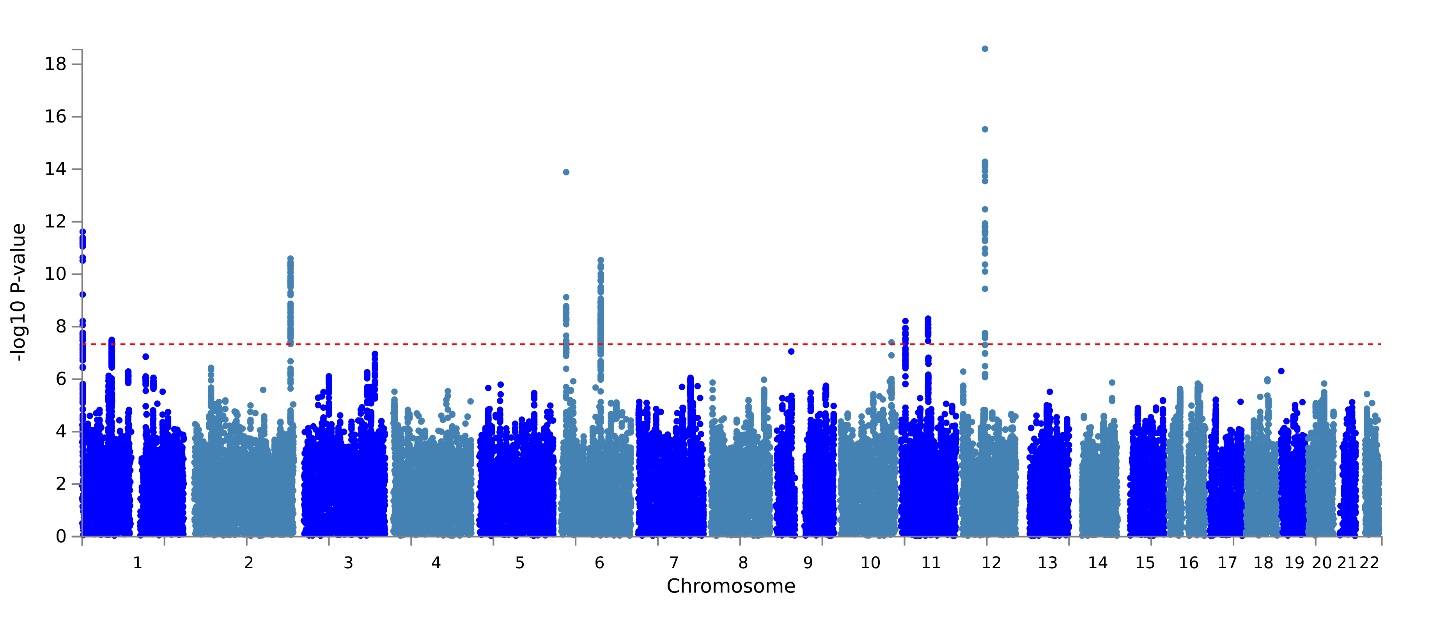

The red dashed line indicates the genome-wide significance threshold (*p* = 5×10⁻⁸).

**Extended Data Fig. 3 | Manhattan plot of migraine genome-wide association results in African ancestry participants from the All of Us Research Program (8,332 cases; 53,348 controls).**
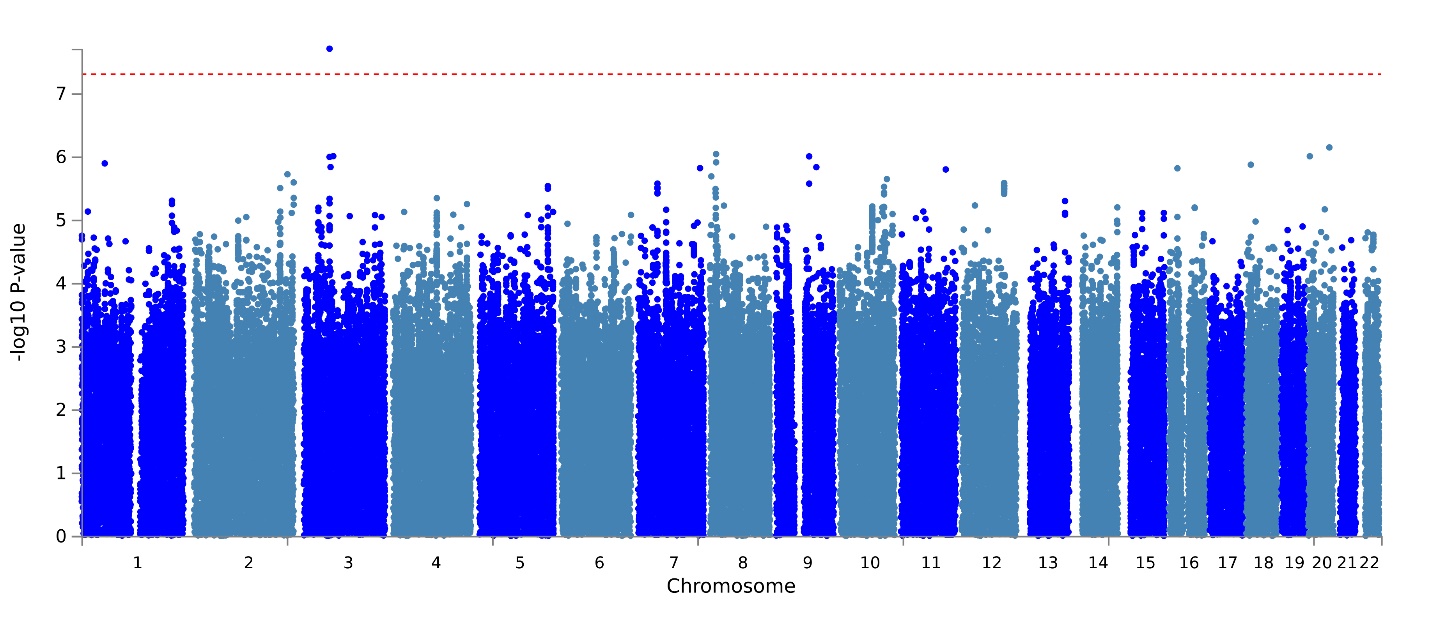

The red dashed line indicates the genome-wide significance threshold (*p* = 5×10⁻⁸).

**Extended Data Fig. 4 | Manhattan plot of migraine genome-wide association results in European ancestry participants from the Yale-Penn cohort (822 cases; 4,569 controls).**
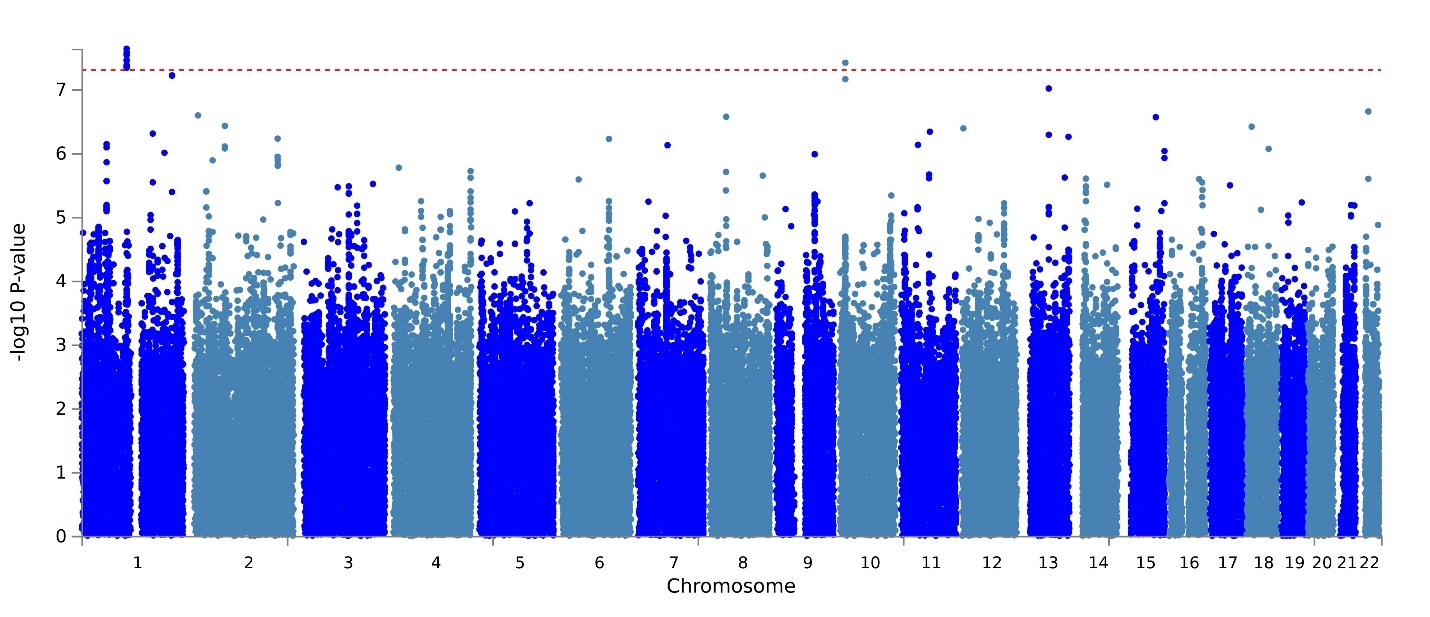

The red dashed line indicates the genome-wide significance threshold (*p* = 5×10⁻⁸).

**Extended Data Fig. 5 | Manhattan plot of migraine genome-wide association results in African ancestry participants from the Yale-Penn cohort (833 cases; 5,081 controls).**
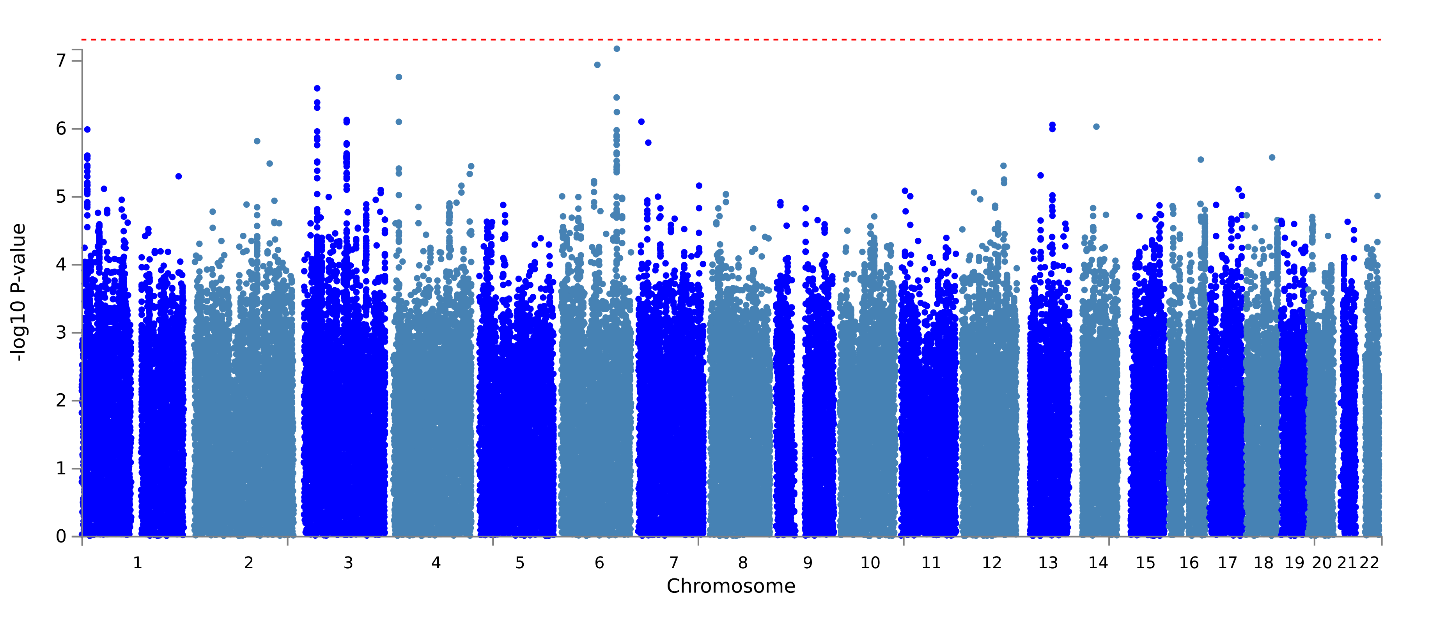

The red dashed line indicates the genome-wide significance threshold (*p* = 5×10⁻⁸).

**Extended Data Fig. 6 | Manhattan plot of the African ancestry migraine meta-analysis.**
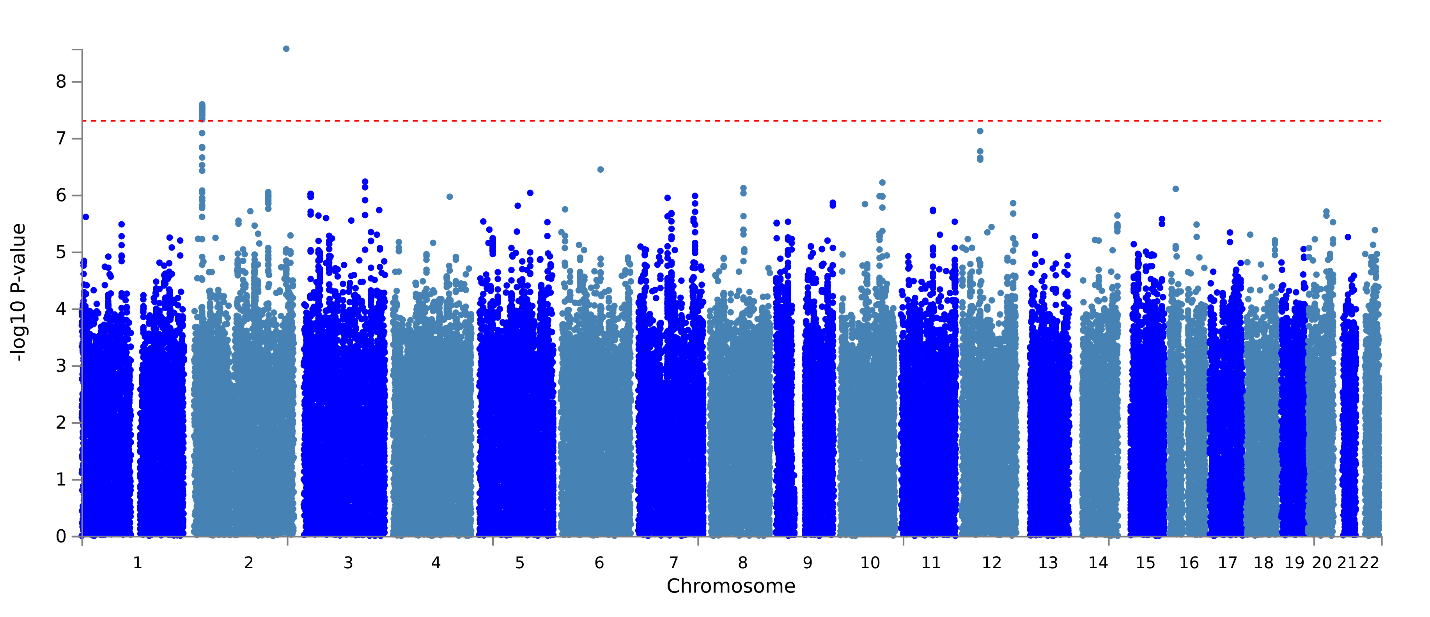

The red dashed line indicates the genome-wide significance threshold (*p* = 5×10⁻⁸).

**Extended Data Fig. 7 | Manhattan plot of the trans-ancestry migraine meta-analysis.**
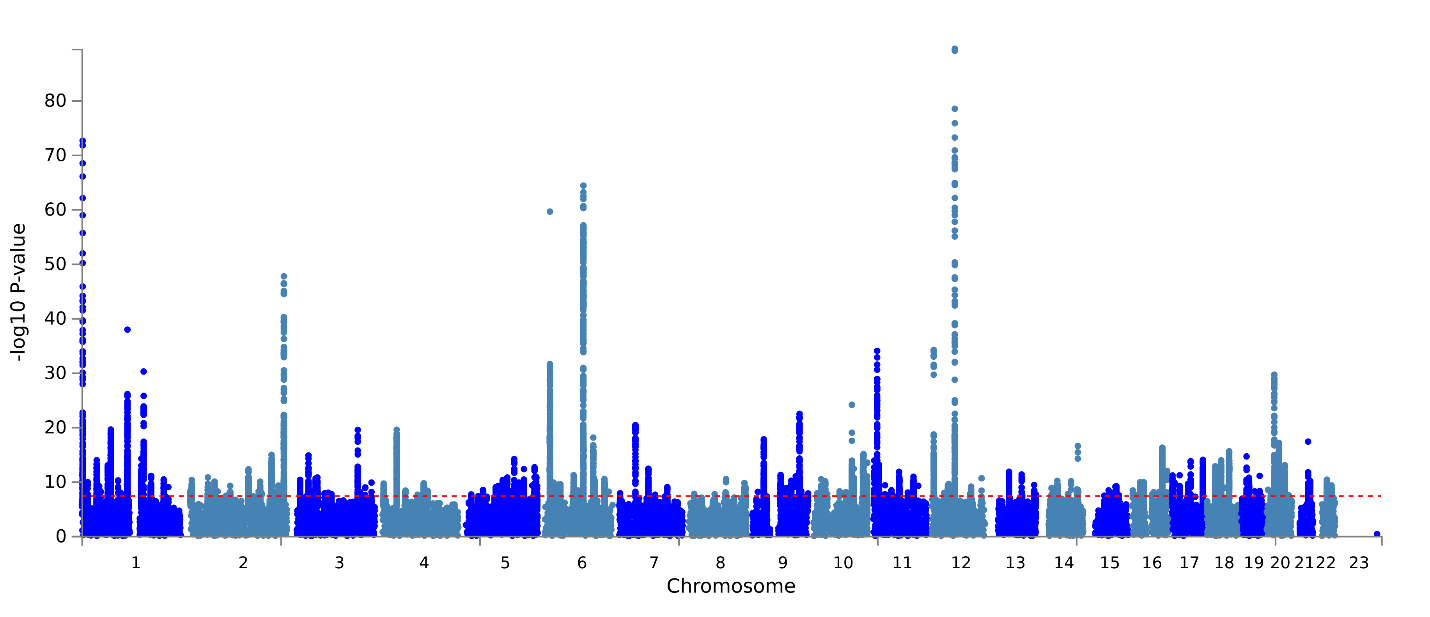

The red dashed line indicates the genome-wide significance threshold (*p* = 5×10⁻⁸).

**Extended Data Fig. 8 | Tissue-specific gene-expression enrichment for the European ancestry migraine meta-analysis using FUMA GENE2FUNC.**
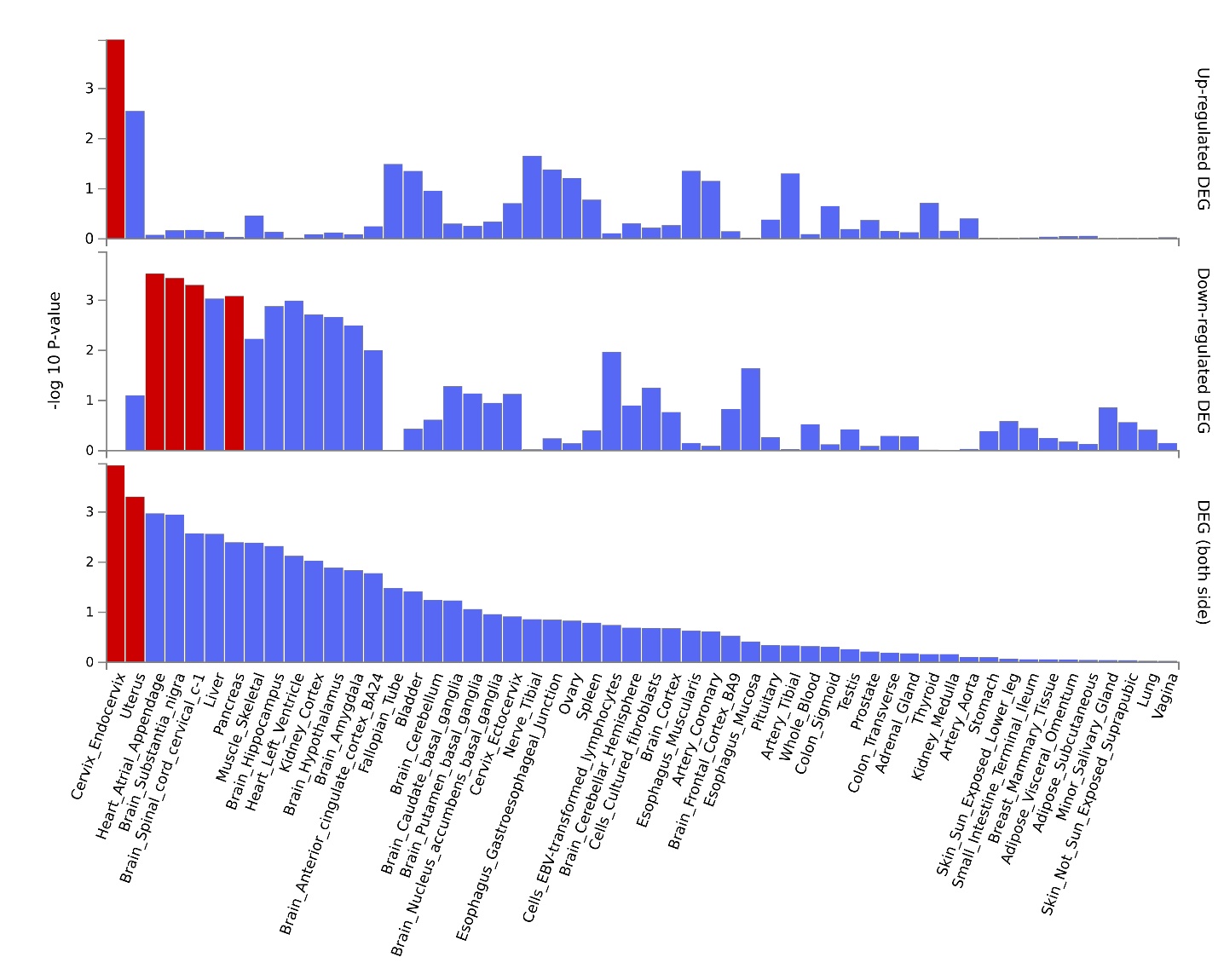

Significantly enriched DEG sets (Bonferroni-corrected *p* < 0.05) are highlighted in red.

**Extended Data Fig. 9 | g:Profiler enrichment results for the TWAS-derived gene set.**

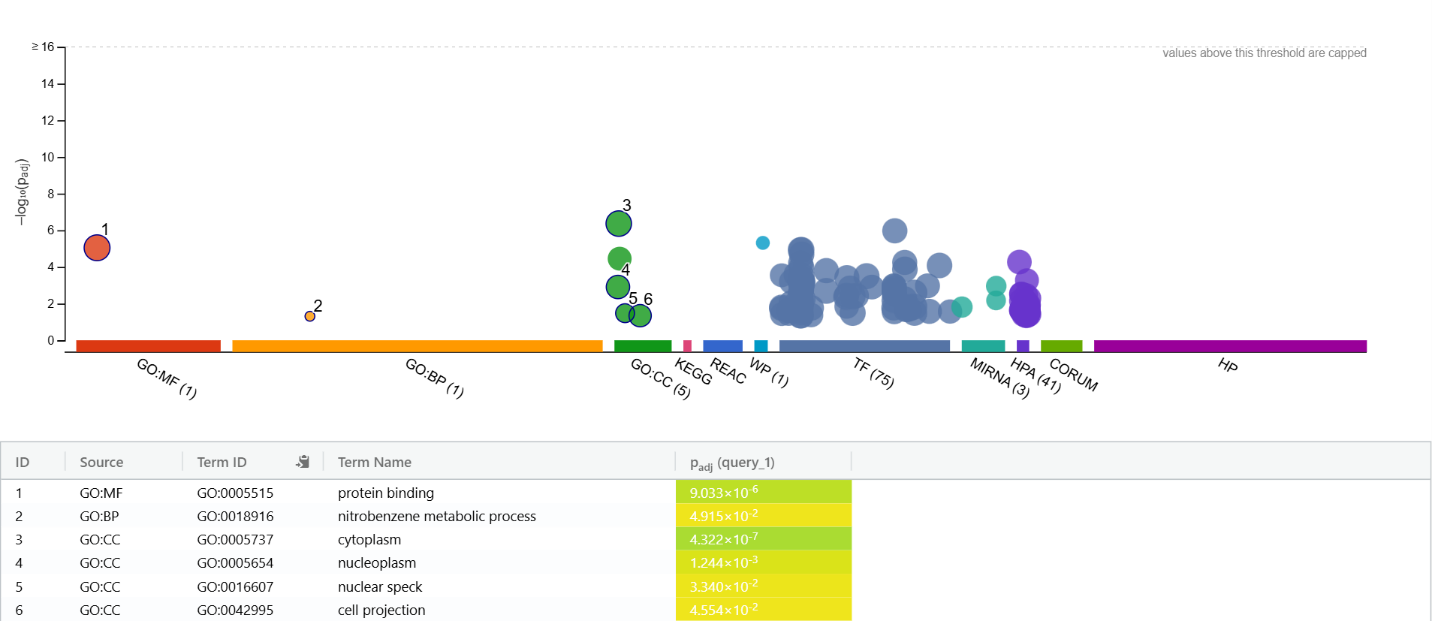
